# Representation of Race and Ethnicity in a Contemporary US Health Cohort: The *All of Us* Research Program

**DOI:** 10.1101/2022.10.26.22281570

**Authors:** Nina Kathiresan, So Mi Jemma Cho, Romit Bhattacharya, Buu Truong, Whitney Hornsby, Pradeep Natarajan

## Abstract

Participation and inclusion in biomedical research remain disproportionate across sociodemographics limiting discoveries in genomic studies and contributing to systemic disparities in healthcare. To alleviate such inequities, the National Institute of Health initiated the *All of Us* Research Program (AoU), a prospective, population-based cohort to identify the root causes and consequences of health outcomes across diverse demographics. We quantify representation of key racial groups in the accruing AoU cohort of US adults aged ≥18 years and compare to their actual representation in the US. Of the 358,705 AoU participants to date, Hispanic or Latino participants were underrepresented by 0.85-fold, non-Hispanic Asian by 0.58-fold, non-Hispanic White by 0.98-fold, and Other by 0.43-fold. Meanwhile, individuals identifying as non-Hispanic Black or African American were overrepresented by 1.88-fold. While AoU representation better mirrors the US demographics compared to other cohorts, recruitment trends for the ongoing AoU underscore the need to further tailor participation initiatives for diverse populations.

## Introduction

Historically, participation and inclusion in biomedical research has been disproportionate across sex, gender, age, race, ethnicity, geography, and socioenvironmental characteristics contributing to systemic disparities in healthcare.^1^ Consequently, clinical trials and epidemiological studies often require oversampling or recalibration to maximize generalizability and are interpreted in limited contexts.^2,3^ However, rigorous statistical approaches incompletely capture substantial heterogeneity in healthcare access, systemic inequities, environmental exposures, cultural practices, individual histories, and lifestyle factors across sociodemographics.^4,5^ Notably, race and ethnicity assessments may provide novel insights into the complex interactions of multidimensional risk factors on health outcomes.^6^

To help address such inequities, the National Institute of Health initiated the *All of Us* Research Program (AoU) ($1.5 billion USD over 10 years), a prospective, population-based cohort that explicitly aims to advance biomedical research among >1 million adults across diverse demographics.^7^ While existing studies have elucidated important findings in genomics, pathophysiology, and epidemiology, they are generally overrepresented for non-Hispanic White individuals. In contrast, a core value of AoU is diversity and inclusivity with the intended recruitment of underrepresented communities, including by race, ethnicity, age, sex, gender identity, sexual orientation, disability status, access to healthcare, wealth, educational attainment, and geography.^7^

Since AoU is a flagship ongoing US federally-funded cohort, characterizing initial representation may inform AoU and related efforts. Thus, we sought to quantify the nationwide- and state-level representations of key racial groups in AoU compared to their actual representation in the US Census.

## Results

### Participant characteristics

Among the 358,705 US adults in the AoU cohort, (58,488) 16.3% identified as Hispanic or Latino, (12,710) 3.5% non-Hispanic Asian, (73,348) 20.5% non-Hispanic Black or African American, (205,457) 57.3% non-Hispanic White, and (8702) 2.4% Other race including individuals reporting multiple categories, respectively (**Table 1**). The racial proportion of the AoU significantly differed from that of the referent US population (ACS) at both nationwide and state levels (**Figure 1, Figures S1&S2**, and **Tables S1-S6**).

**Table 1.** Racial and ethnic proportion in the US and *All of Us*

| Race and ethnicity | US |  | *All of Us* |  | Absolute difference (95% CI)<sup>a</sup> |  |
| --- | --- | --- | --- | --- | --- | --- |
|  | Count | Proportion | Count | Proportion | (*All of Us* - US) | OR (95% CI) |
| Total | 331,449,281 |  | 358,705 |  |  |  |
| Hispanic or Latino | 62,080,044 | 18.73% | 58,488 | 16.31% | -2.42% (-2.54, -2.30) | 0.85 (0.84, 0.85) |
| Non-Hispanic Asian | 19,618,719 | 5.92% | 12,710 | 3.54% | -2.38% (-2.44, -2.32) | 0.58 (0.57, 0.59) |
| Non-Hispanic Black or African American | 39,940,338 | 12.05% | 73,348 | 20.45% | 8.40% (8.27, 8.53) | 1.88 (1.86, 1.89) |
| Non-Hispanic White | 191,697,647 | 57.84% | 205,457 | 57.28% | -0.56% (-0.72, -0.40) | 0.98 (0.97, 0.98) |
| Other including multiracial | 18,112,533 | 5.46% | 8,702 | 2.43% | -3.03% (-3.09, -2.99) | 0.43 (0.42, 0.44) |

Abbreviations: CI, confidence interval; OR, odds ratio
<sup>a</sup>Derived from the 2-sample test for equality of proportions

**Figure 1.**
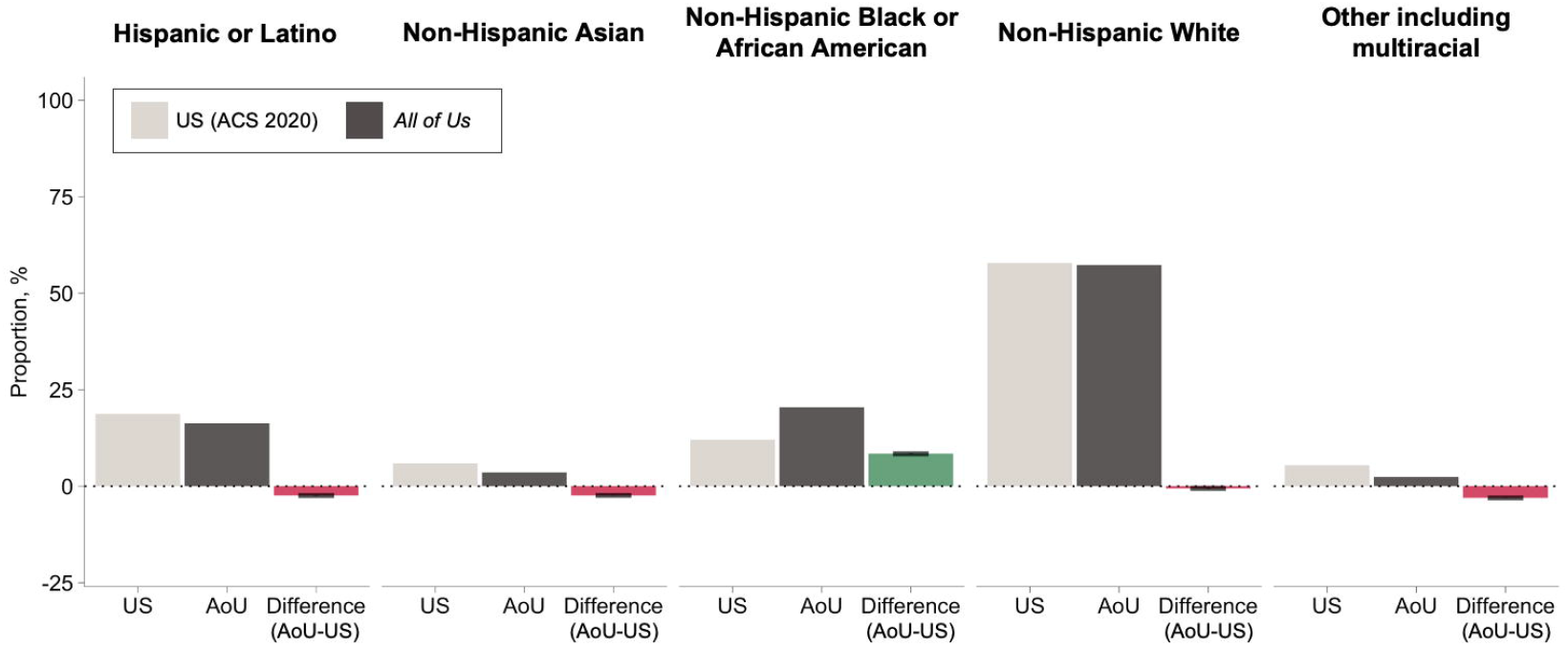
Comparison of key race/ethnicity group prevalences between the US and *All of Us*. The prevalences of self-reported race/ethnicity groups reported in both *All of Us* and the U.S. Census across the full datasets as well as differences are presented. Abbreviation: AoU, *All of Us* ^a^Absolute difference (95% confidence interval) is calculated using the 2-sample test for equality of proportions.

Additionally, 96,268 of 358,705 (26.84%) AoU participants had available genetic ancestry estimation (**Figure S3, Table S7**). Of them, 23.4% (22,501) were classified as African, 2.2% (2094) East Asian, 48.8% (47,022) European, 15.4% (14,845) Latino/Admixed American, 0.2% (161) Middle Eastern, 9.0% (8690) Other, and 0.99% (955) South Asian, respectively. The extent of genetic admixture varied across self-reported race and ethnicity categories (**Figure 3, Figure S4**). Notably, self-reported Hispanic or Latino individuals displayed substantial genetic heterogeneity in contrast to higher concordance observed in the Non-Hispanic Black or African American category.

### Representation of race and ethnicity in the AoU

The greatest relative underrepresentation was observed in Other or multiracial category in the AoU cohort (2.4%) relative to the US (5.5%) (absolute difference, -3.03% [95% CI -3.09, -2.99]) by 0.43-fold, which extended across nearly every state (**Figure 2, Table S6**). Furthermore, non-Hispanic Asian participants were 0.58-fold likely (absolute difference, -2.38% [-2.44, -2.32]) to be represented in AoU. Individuals identifying as Hispanic or Latino, the second largest race group in the US, were 0.85-fold underrepresented in the AoU cohort (AoU, 16.3% vs US, 18.7%) and most pronounced in Puerto Rico by 30.7% (**Table S2**).

**Figure 2.**
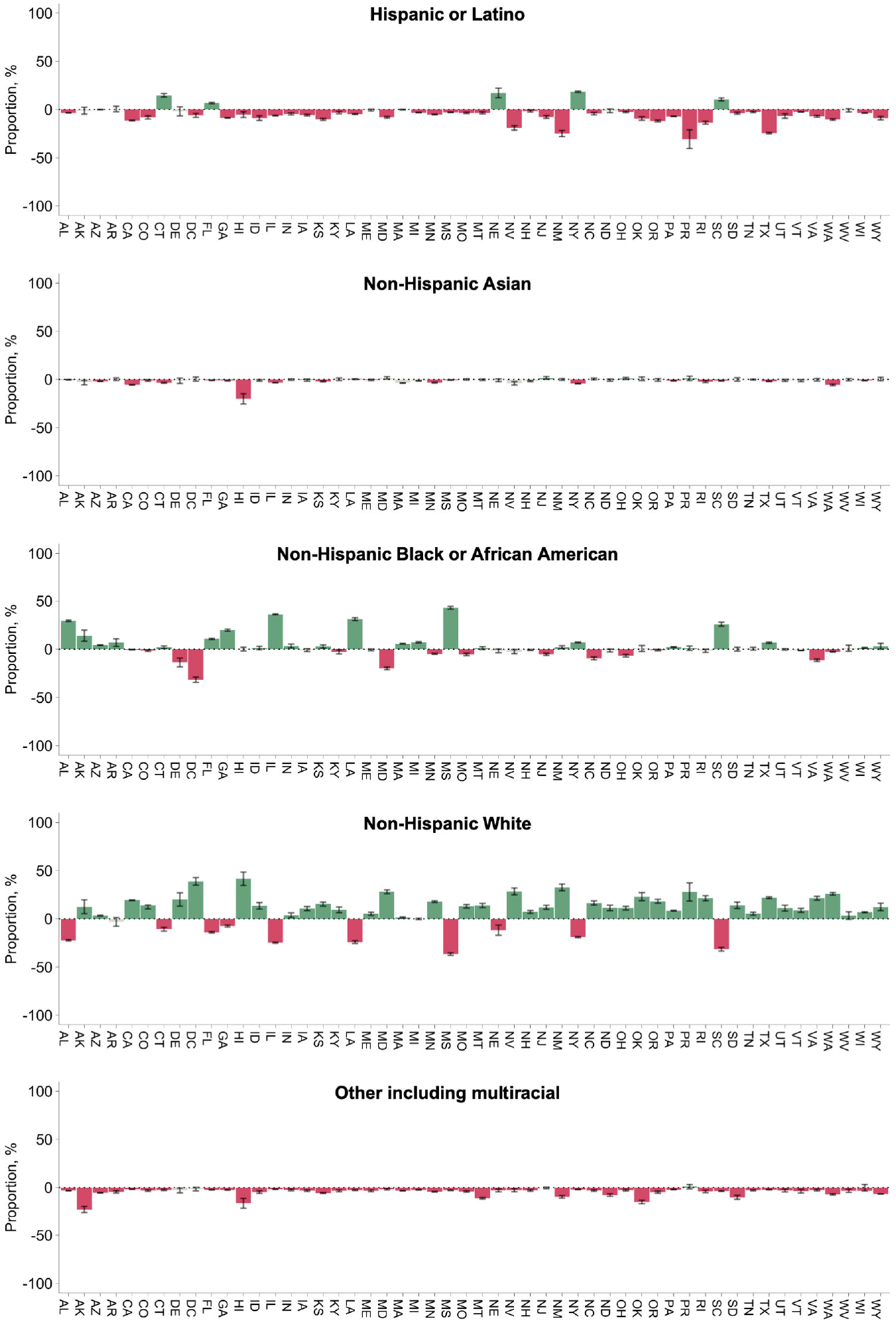
Differences in key racial representation between the US and *All of Us* by state. The prevalence differences for each self-reported race/ethnicity group between *All of Us* and the U.S. Census by state are presented. ^a^Red bar indicates statistically significant underrepresentation in *All of Us*; green bar indicates significant overrepresentation in *All of Us*; gray bar indicates statistically nonsignificant difference. ^b^Absolute difference (95% confidence interval) is calculated using the 2-sample test for equality of proportions.

**Figure 3.**
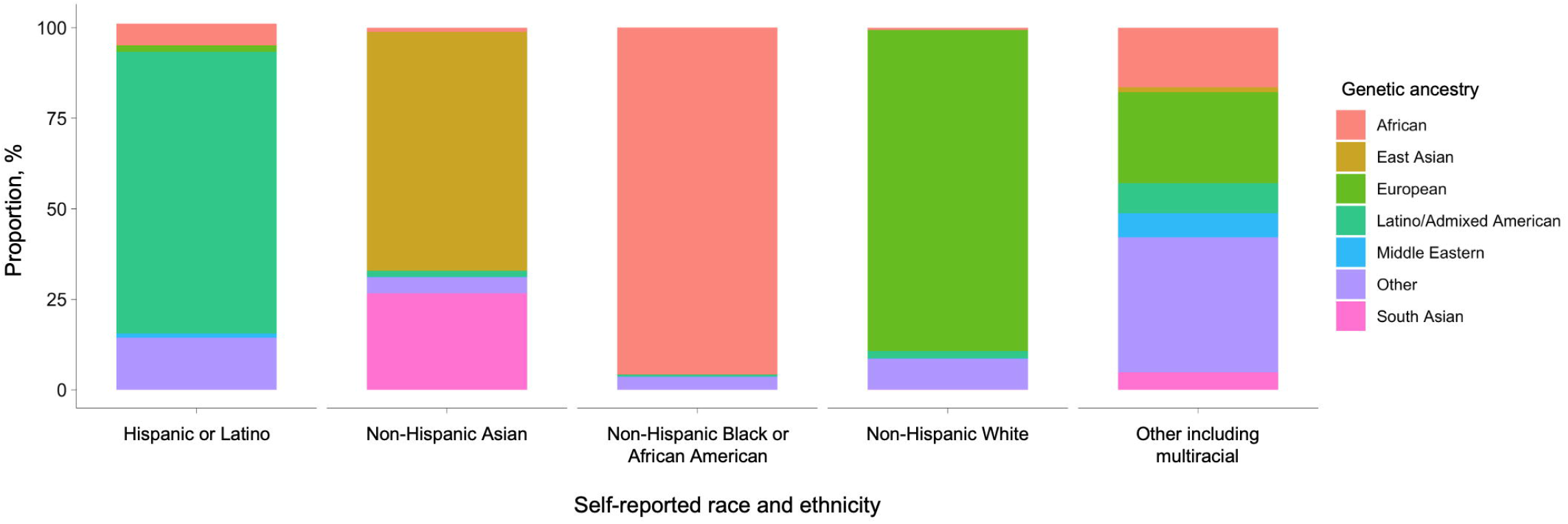
Heterogeneity in genetic ancestry within self-reported race and ethnicity in *All of Us*. ^a^All values are presented as proportion within each self-reported race and ethnicity category. ^b^Color codings reflect genetic ancestry inferred from 96,268 of 358,705 participants with whole genome sequencing data. ^c^Genetic ancestry categories are consistent with the gnomAD, Human Genome Diversity Project, and 1000 Genomes classifications. ^d^Individuals were binned into genetically-inferred ancestry groups. Concordance with self-reported race and ethnicity was evaluated. The present plot compares these two metrics.

Non-Hispanic White individuals accounted for the greatest absolute participation in the AoU with generally consistent dominance across all states with marginal relative underrepresentation by 0.98-fold (absolute difference, -0.56% [-0.72, -0.40]) to the US (**Table S5**). Only non-Hispanic Black or African American individuals were overrepresented in the AoU by 1.88-fold and by absolute scale, 8.40% (AoU, 20.5% vs US, 12.1%), with similar trends across most states (**Table S4**).

## Discussion

In this contemporary nationwide biomedical cohort, we observed underrepresentation of Hispanic or Latino, non-Hispanic Asian, and Other multiracial populations. On the other hand, non-Hispanic Black or African American population, historically regarded as understudied,^1^ accounted for the second largest proportion and was overrepresented across most states. Furthermore, notable genetically-inferred continental ancestry heterogeneity was observed within each self-reported race category. These findings underscore the need to further improve recruitment and participation initiatives for diverse populations for the ongoing AoU Research Program as well as improved tools to better capture demographic diversity.

Equitable sociodemographic representation in biomedical datasets is imperative to translate research investments into both generalizable and targeted advances. Previous literature has established a gradient of health risk attributable to features closely linked to individual and perceived racial and ethnic identity, often manifested through socioeconomic injustice, deprivation, environmental inequities, psychosocial resilience, differential access to healthcare, and political exclusion.^8^ Even within a given racial and ethnic category, each subpopulation embodies a unique product of risk factors and experiences shaped by migration and culture.^9^ However, the consistent underrepresentation of non-White individuals in US biomedical research continues to exacerbate limited understanding of root causes of health outcomes. While initial AoU recruitment trends are encouraging for non-Hispanic Black individuals, other groups remain starkly underrepresented despite the explicit goals of AoU. Towards promoting equity, initiatives should not merely target proportionate representation but ideally overrepresentation of historically underrepresented communities and adequately redistribute investments.

We also observed that heterogeneity in genetic ancestry varied differentially across self-reported racial and ethnic categories. Contemporary epidemiological studies often interchangeably use race, ethnicity, and ancestry without clear distinctions of measured domains.^10^ The principal components for population stratification are based on allele frequencies and co-occurrences observed in groups and have largely been used to minimize the possibility of spurious genetic associations in discovery studies.^11^ On the other hand, self-identified race and ethnicity is a subjective interpretation of a complex combination of both genetic and non-genetic information, such as culture, societal construct, and physical appearance^12^ otherwise uncaptured at an individual genomics level. A recent multiethnic comparative study^13^ illustrated the differential genome-wide association effect with coronary artery disease even within a single ancestral population, concluding that aggregating i.e., Hispanics into a singular category may overlook differences within a subpopulation. To achieve more precise and equitable disease prediction, prevention, and intervention, future biomedical studies warrant more comprehensive and sensitive approaches to probe into underlying variability in genetics and social and physical environments.

### Limitations

Our findings should be interpreted in the context of potential limitations. Responses on sociodemographics were restricted to questions included in the AoU survey. Racial and ethnic sub-classifications or excluded categories may reveal further heterogeneity. The state-specific results are subjected to change pending immigration or emigration. Nevertheless, the current study adhered to classification consistent with the most recent US Census Bureau scheme.

### Conclusion

Historical underrepresentation of Hispanic or Latino, non-Hispanic Asian, and multiracial populations persist in the largest ongoing single federally funded biomedical research cohort in the US. Given ongoing recruitment toward greater than 1 million, reallocation of resources toward more equitable participation is needed to mitigate the downstream risks of further exacerbating disparities in healthcare.

## Supporting information

STAR Methods table

## Data Availability

American Community Survey 2020 aggregate data are publicly available at https://www.census.gov/programs-surveys/acs and *All of Us* controlled access individual-level data are available at https://allofus.nih.gov. All original code and any additional information required to reanalyze the data reported in this paper are available from the lead contact upon reasonable request.

https://allofus.nih.gov

## Acknowledgments

Dr. Cho is supported by a grant of the Korea Health Technology R&D Project through the Korea Health Industry Development Institute (KHIDI), funded by the Ministry of Health & Welfare, Republic of Korea (grant no.: HI19C1330). Dr. Bhattacharya is supported by the John S. LaDue Memorial Fellowship in Cardiovascular Research. Dr. Natarajan is supported by a grant from the National Human Genetics Research Institute (U01HG011719) and Massachusetts General Hospital (Paul and Phyllis Fireman Endowed Chair in Vascular Medicine). The funders/sponsors had no role in the design and conduct of the study; collection, management, analysis, and interpretation of the data; preparation, review, or approval of the manuscript; and decision to submit the manuscript for publication.

## Author contributions

Conceptualization, RB, PN; Methodology, SMJC, PN; Software, SMJC; Validation, SMJC; Formal Analysis, SMJC, BT; Investigation, NK, SMJC, PN; Resources, PN; Data Curation, SMJC, BT; Writing-original draft, NK, SMJC; Writing-review & editing, NK, SMJC, RB, BT, WH, PN; Visualization, SMJC, BT; Supervision, PN; Project administration, RB, PN; Funding acquisition, PN

## STAR Methods

### RESOURCE AVAILABILITY

#### Lead contact

Further information and requests for resources should be directed to and will be fulfilled by the lead contact, Pradeep Natarajan.

#### Materials availability

This study did not generate new unique reagents.

#### Data and code availability

The All of Us Research Program (AoU) data is publicly available to researchers upon application at https://www.researchallofus.org/register/. The application requires institutional Data Use and Registration Agreement to ensure data security; agreement and adherence to the data Use Code of Conduct; verification of identity using LOGIN.GOV (https://login.gov/what-is-login/); completion of the mandatory AoU research training; familiarity with the Research Workbench tools as well as data access tiers contingent upon use of aggregate-versus individual-level data and; details on proposed research objectives, scope, methods, and expected clinical/population implications resulting from the study. The American Community Survey (ACS) data is publicly available at https://data.census.gov/cedsci/. The nationwide and state-level distributions of key racial/ethnic groups can be viewed by specifying specific race aggregation or geography options.

All original code and any additional information required to reanalyze the data reported in this paper are available from the lead contact upon reasonable request.

### METHOD DETAILS

#### Self-reported race and ethnicity in ACS and AoU

The Massachusetts General Hospital institutional review board approved the secondary use of the ACS 2020 (2021P002212) and AoU Research Program data (2020P001737). Informed consent for American Community Survey (ACS) was waived because it is routinely collected and deidentified data. All participants of the AoU provided informed consent. Reporting follows the Strengthening the Reporting of Observational Studies in Epidemiology (STROBE) guidelines.

The ACS is an annually aggregated household survey conducted by the US Census Bureau, which collects population-level sociodemographic, economic, and residential metrics.^14^ The ACS is administered to a random rolling sampling of 295 000 addresses monthly via the internet, mail, telephone, and in-person interviews.^14^ With ongoing recruitment since 2017, AoU is a nationwide, prospective volunteer cohort study aiming to study health outcomes, risk factors, novel biomarkers, and social and behavioral determinants of health across US communities.^7^ AoU participants aged ≥18 years who consented to sociodemographic survey, anthropometry, biospecimen collection, and linkage to healthcare utilization data were included. Details of AoU were previously described.^15^ The present analysis used AoU v6, which comprised 358 705 participants with available race and residential state data collected between May 31, 2017 and January 1, 2022. ACS aggregate data are publicly available at https://www.census.gov/programs-surveys/acs and AoU controlled access individual-level data are available at https://allofus.nih.gov. In accordance with the US Census Bureau, we identified five major racial categories, which include Hispanic or Latino, non-Hispanic Asian, non-Hispanic Black or African American, non-Hispanic White, and Other including non-Hispanic individuals reporting multiple races.

#### Quality control of AoU genotypic data

The present analysis relied on Q2 2022 release of genomic data made available in the AoU Researcher Workbench on June 22th, 2022, which contains 165,127 array samples and 98,590 whole genome sequencing (WGS) samples. The AoU Genome Centers and Data Research Center have performed imputation of genotypic data and extensive quality control to minimize batch effects, to prevent sample swaps or contamination, and to filter variants.^16^ Briefly, genotypes were processed using the Illumina Global Diversity Array (Illumina, Inc., San Diego, California, USA).^16^ At batch level, the samples with log-likelihood ratio >-3 concordance between the array and WGS, matched self-reported sex assigned at birth and genotyped sex, >98% call rate, and <3% cross-individual contamination rate were accessioned.^16^

Likewise, all centers applied the same protocols and singularly utilized the Illumina, Inc. (San Diego, California, USA) tools for library construction (PCR Free Kapa HyperPrep), quantification (Illumina Kapa DNA Quantification Kit), sequencing (NovaSeq 6000), and analysis (DRAGEN Platform v3.4.12) for WGS.^17^ The GRCh38DH reference genome was used for alignment (available at: ftp://ftp.1000genomes.ebi.ac.uk/vol1/ftp/technical/reference/GRCh38_reference_genome/).^18^ Quality control on joint callsets has been performed, including hard threshold flagging, population outlier flagging, hard threshold filtering, Allele-Specific Variant Quality Score Recalibration,^19^ and sensitivity and precision evaluation.

#### Inferring of genomic ancestry in the AoU cohort

For all 98,590 WGS samples, the AoU readily classified and provided ancestry consistent with the gnomAD,^20^ Human Genome Diversity Project,^21^ and 1000 Genomes classification^22^: i) African; ii) Latino/Native American/Admixed American; iii) East Asian; iv) Middle Eastern; v) European; vi) Other; and vi) South Asian. Random forest classifier^23^ method was trained on the Human Genome Diversity Project and 1kg samples variants on chromosomes 20 and 21 obtained from gnomAD.^24^ The first 16 principal components were generated using the *hwe_normalized_pca* method in Hail^25^ at high-quality variant sites,^26^ defined as i) inclusion of autosomal, bi-allelic single nucleotide variants only; ii) allele frequency >0.1%; iii) call rate >99%; iv) linkage disequilibrium-pruned with a cutoff of r^2^=0.1.^16^ Individuals with probability of predicted ancestry ≤90% by the random forest model was categorized as “Other.” The AoU reports the concordance rate of 0.877 between self-reported ethnicity and the ancestry predictions after excluding participants without response or selecting “prefer not to answer” or “skip” in the questionnaire.^16^

### QUANTIFICATION AND STATISTICAL ANALYSIS

We calculated the proportion of each race group by diving the total count of 331,449,281 for ACS and 358,705 for AoU, respectively. Per the AoU Data and Statistics Dissemination Policy, participant counts ≤20 are masked with an asterisk.^15^ We performed chi-squared tests to test for relative differences in racial distributions in the US and, separately, by state. Then, we used 2-sample tests for equality of proportions to estimate absolute differences for each race between ACS and AoU. To depict in relative scale, we also presented the representation differences in odds ratio (OR) for each race group. Statistically significant under- or over-representation of each race category in AoU was determined with nonoverlapping 95% confidence intervals. As a secondary analysis, among AoU participants with WGS data, we estimated the genetic diversity within each self-reported race and ethnicity category. All statistical tests were two-sided, and statistical significance was set at *P* <0.05. All analyses were performed using R version 4.1.0 (R Foundation for Statistical Computing, Vienna, Austria).

