## Supplemental File for "Representation of Race and Ethnicity in a Contemporary US Health Cohort: The *All of Us* Research Program"

**Table S1.** Differences in racial and ethnic representation between the US and *All of Us* by state

**Table S2.** Differences in Hispanic or Latino representation between the US and *All of Us*

**Table S3.** Differences in non-Hispanic Asian representation between the US and *All of Us*

**Table S4.** Differences in non-Hispanic Black or African American representation between the US and *All of Us*

**Table S5.** Differences in non-Hispanic White representation between the US and *All of Us*

**Table S6.** Differences in Other race and ethnicity representation between the US and *All of Us*

**Table S7.** Diversity of genetic ancestry within each self-reported racial and ethnic category in *All of Us*

**Figure S1.** Comparison of racial and ethnic proportions between the US and *All of Us*

**Figure S2.** Comparison of racial and ethnic proportions between the US and *All of Us* by state

**Figure S3.** Principal component plot of *All of Us* participants

**Figure S4.** Heterogeneity in genetic ancestry within self-reported racial and ethnic category in *All of Us*

| Table S1. Differences in racial and ethnic representation between the US and *All of Us* by state | | | | | |  |  |  |
| --- | --- | --- | --- | --- | --- | --- | --- | --- |
|  |  | US | |  | *All of Us* | |  |  |
| State | Race and ethnicity | Count | Proportion, % |  | Count | Proportion, % |  | *P* value^b^ |
| Alabama | Total | 5,024,279 |  |  | 21,640 |  |  | <0.001 |
|  | Hispanic or Latino | 264,047 | 5.26 |  | 382 | 1.77 |  |  |
|  | Non-Hispanic Asian | 75,918 | 1.51 |  | 236 | 1.09 |  |  |
|  | Non-Hispanic Black or African American | 1,288,159 | 25.64 |  | 11,926 | 55.11 |  |  |
|  | Non-Hispanic White | 3,171,351 | 63.12 |  | 8,803 | 40.68 |  |  |
|  | Other including multiracial | 224,804 | 4.47 |  | 293 | 1.35 |  |  |
| Alaska | Total | 733,391 |  |  | * |  |  | <0.001 |
|  | Hispanic or Latino | 49,824 | 6.79 |  | * | 5.63 |  |  |
|  | Non-Hispanic Asian | 43,449 | 5.92 |  | * | 3.13 |  |  |
|  | Non-Hispanic Black or African American | 20,731 | 2.83 |  | * | 16.88 |  |  |
|  | Non-Hispanic White | 421,758 | 57.51 |  | * | 70.00 |  |  |
|  | Other including multiracial | 197,629 | 26.95 |  | * | 4.38 |  |  |
| Arizona | Total | 7,151,502 |  |  | 44,445 |  |  | <0.001 |
|  | Hispanic or Latino | 2,192,253 | 30.65 |  | 13,486 | 30.34 |  |  |
|  | Non-Hispanic Asian | 248,837 | 3.48 |  | 690 | 1.55 |  |  |
|  | Non-Hispanic Black or African American | 317,161 | 4.43 |  | 3,865 | 8.70 |  |  |
|  | Non-Hispanic White | 3,816,547 | 53.37 |  | 25,209 | 56.72 |  |  |
|  | Other including multiracial | 576,704 | 8.06 |  | 1,195 | 2.69 |  |  |
| Arkansas | Total | 3,011,524 |  |  | * |  |  | <0.001 |
|  | Hispanic or Latino | 256,847 | 8.53 |  | * | 9.09 |  |  |
|  | Non-Hispanic Asian | 51,210 | 1.70 |  | * | 1.91 |  |  |
|  | Non-Hispanic Black or African American | 449,884 | 14.94 |  | * | 21.77 |  |  |
|  | Non-Hispanic White | 2,063,550 | 68.52 |  | * | 65.31 |  |  |
|  | Other including multiracial | 190,033 | 6.31 |  | * | 1.91 |  |  |
| California | Total | 39,538,223 |  |  | 47,842 |  |  | <0.001 |
|  | Hispanic or Latino | 15,579,652 | 39.40 |  | 13,400 | 28.01 |  |  |
|  | Non-Hispanic Asian | 5,978,795 | 15.12 |  | 4,553 | 9.52 |  |  |
|  | Non-Hispanic Black or African American | 2,119,286 | 5.36 |  | 2,289 | 4.78 |  |  |
|  | Non-Hispanic White | 13,714,587 | 34.69 |  | 25,816 | 53.96 |  |  |
|  | Other including multiracial | 2,145,903 | 5.43 |  | 1,784 | 3.73 |  |  |
| Colorado | Total | 5,773,714 |  |  | 1,601 |  |  | <0.001 |
|  | Hispanic or Latino | 1,263,390 | 21.88 |  | 220 | 13.74 |  |  |
|  | Non-Hispanic Asian | 195,220 | 3.38 |  | 35 | 2.19 |  |  |
|  | Non-Hispanic Black or African American | 221,310 | 3.83 |  | 38 | 2.37 |  |  |
|  | Non-Hispanic White | 3,760,663 | 65.13 |  | 1,266 | 79.08 |  |  |
|  | Other including multiracial | 333,131 | 5.77 |  | 42 | 2.62 |  |  |
| Connecticut | Total | 3,605,944 |  |  | 2,464 |  |  | <0.001 |
|  | Hispanic or Latino | 623,293 | 17.29 |  | 791 | 32.10 |  |  |
|  | Non-Hispanic Asian | 170,459 | 4.73 |  | 31 | 1.26 |  |  |
|  | Non-Hispanic Black or African American | 360,937 | 10.01 |  | 296 | 12.01 |  |  |
|  | Non-Hispanic White | 2,279,232 | 63.21 |  | 1,289 | 52.31 |  |  |
|  | Other including multiracial | 172,023 | 4.77 |  | 57 | 2.31 |  |  |
| Delaware | Total | 989,948 |  |  | * |  |  | <0.001 |
|  | Hispanic or Latino | 104,290 | 10.53 |  | * | 8.57 |  |  |
|  | Non-Hispanic Asian | 42,398 | 4.28 |  | * | 2.86 |  |  |
|  | Non-Hispanic Black or African American | 212,960 | 21.51 |  | * | 7.86 |  |  |
|  | Non-Hispanic White | 579,851 | 58.57 |  | * | 78.57 |  |  |
|  | Other including multiracial | 50,449 | 5.10 |  | * | 2.14 |  |  |
| District of Columbia | Total | 689,545 |  |  | * |  |  | <0.001 |
|  | Hispanic or Latino | 77,652 | 11.26 |  | * | 5.26 |  |  |
|  | Non-Hispanic Asian | 33,192 | 4.81 |  | * | 5.26 |  |  |
|  | Non-Hispanic Black or African American | 282,066 | 40.91 |  | * | 9.38 |  |  |
|  | Non-Hispanic White | 261,771 | 37.96 |  | * | 76.66 |  |  |
|  | Other including multiracial | 34,864 | 5.06 |  | * | 3.43 |  |  |
| Florida | Total | 21,538,187 |  |  | 17,123 |  |  | <0.001 |
|  | Hispanic or Latino | 5,697,240 | 26.45 |  | 5,697 | 33.27 |  |  |
|  | Non-Hispanic Asian | 629,626 | 2.92 |  | 339 | 1.98 |  |  |
|  | Non-Hispanic Black or African American | 3,127,052 | 14.52 |  | 4,297 | 25.09 |  |  |
|  | Non-Hispanic White | 11,100,503 | 51.54 |  | 6,409 | 37.43 |  |  |
|  | Other including multiracial | 983,766 | 4.57 |  | 381 | 2.23 |  |  |
| Georgia | Total | 10,711,908 |  |  | 9,018 |  |  | <0.001 |
|  | Hispanic or Latino | 1,123,457 | 10.49 |  | 165 | 1.83 |  |  |
|  | Non-Hispanic Asian | 475,680 | 4.44 |  | 289 | 3.20 |  |  |
|  | Non-Hispanic Black or African American | 3,278,119 | 30.60 |  | 4,557 | 50.53 |  |  |
|  | Non-Hispanic White | 5,362,156 | 50.06 |  | 3,829 | 42.46 |  |  |
|  | Other including multiracial | 472,496 | 4.41 |  | 178 | 1.97 |  |  |
| Hawaii | Total | 1,455,271 |  |  | * |  |  | <0.001 |
|  | Hispanic or Latino | 138,923 | 9.55 |  | * | 4.35 |  |  |
|  | Non-Hispanic Asian | 531,558 | 36.53 |  | * | 16.30 |  |  |
|  | Non-Hispanic Black or African American | 21,877 | 1.50 |  | * | 1.63 |  |  |
|  | Non-Hispanic White | 314,365 | 21.60 |  | * | 63.04 |  |  |
|  | Other including multiracial | 448,548 | 30.82 |  | * | 14.67 |  |  |
| Idaho | Total | 1,839,106 |  |  | * |  |  | <0.001 |
|  | Hispanic or Latino | 239,407 | 13.02 |  | * | 4.08 |  |  |
|  | Non-Hispanic Asian | 26,036 | 1.42 |  | * | 0.41 |  |  |
|  | Non-Hispanic Black or African American | 14,785 | 0.80 |  | * | 2.04 |  |  |
|  | Non-Hispanic White | 1,450,523 | 78.87 |  | * | 92.24 |  |  |
|  | Other including multiracial | 108,355 | 5.89 |  | * | 1.22 |  |  |
| Illinois | Total | 12,812,508 |  |  | 32,636 |  |  | <0.001 |
|  | Hispanic or Latino | 2,337,410 | 18.24 |  | 3,882 | 11.89 |  |  |
|  | Non-Hispanic Asian | 747,280 | 5.83 |  | 815 | 2.50 |  |  |
|  | Non-Hispanic Black or African American | 1,775,612 | 13.86 |  | 16,357 | 50.12 |  |  |
|  | Non-Hispanic White | 7,472,751 | 58.32 |  | 10,920 | 33.46 |  |  |
|  | Other including multiracial | 479,455 | 3.74 |  | 662 | 2.03 |  |  |
| Indiana | Total | 6,785,528 |  |  | 1,161 |  |  | <0.001 |
|  | Hispanic or Latino | 554,191 | 8.17 |  | 42 | 3.62 |  |  |
|  | Non-Hispanic Asian | 166,651 | 2.46 |  | 27 | 2.33 |  |  |
|  | Non-Hispanic Black or African American | 637,500 | 9.39 |  | 147 | 12.66 |  |  |
|  | Non-Hispanic White | 5,121,004 | 75.47 |  | 920 | 79.24 |  |  |
|  | Other including multiracial | 306,182 | 4.51 |  | 25 | 2.15 |  |  |
| Iowa | Total | 3,190,369 |  |  | * |  |  | <0.001 |
|  | Hispanic or Latino | 215,986 | 6.77 |  | * | 0.94 |  |  |
|  | Non-Hispanic Asian | 75,017 | 2.35 |  | * | 1.69 |  |  |
|  | Non-Hispanic Black or African American | 129,321 | 4.05 |  | * | 3.20 |  |  |
|  | Non-Hispanic White | 2,638,201 | 82.69 |  | * | 93.23 |  |  |
|  | Other including multiracial | 131,844 | 4.13 |  | * | 0.94 |  |  |
| Kansas | Total | 2,937,880 |  |  | * |  |  | <0.001 |
|  | Hispanic or Latino | 382,603 | 13.02 |  | * | 2.74 |  |  |
|  | Non-Hispanic Asian | 85,225 | 2.90 |  | * | 0.81 |  |  |
|  | Non-Hispanic Black or African American | 163,352 | 5.56 |  | * | 8.32 |  |  |
|  | Non-Hispanic White | 2,122,575 | 72.25 |  | * | 87.51 |  |  |
|  | Other including multiracial | 184,125 | 6.27 |  | * | 0.61 |  |  |
| Kentucky | Total | 4,505,836 |  |  | * |  |  | <0.001 |
|  | Hispanic or Latino | 207,854 | 4.61 |  | * | 1.53 |  |  |
|  | Non-Hispanic Asian | 73,843 | 1.64 |  | * | 1.79 |  |  |
|  | Non-Hispanic Black or African American | 357,764 | 7.94 |  | * | 5.12 |  |  |
|  | Non-Hispanic White | 3,664,764 | 81.33 |  | * | 90.54 |  |  |
|  | Other including multiracial | 201,611 | 4.47 |  | * | 1.02 |  |  |
| Louisiana | Total | 4,657,757 |  |  | 3,885 |  |  | <0.001 |
|  | Hispanic or Latino | 322,549 | 6.92 |  | 84 | 2.16 |  |  |
|  | Non-Hispanic Asian | 85,336 | 1.83 |  | 84 | 2.16 |  |  |
|  | Non-Hispanic Black or African American | 1,452,420 | 31.18 |  | 2,425 | 62.42 |  |  |
|  | Non-Hispanic White | 2,596,702 | 55.75 |  | 1,226 | 31.56 |  |  |
|  | Other including multiracial | 200,750 | 4.31 |  | 66 | 1.70 |  |  |
| Maine | Total | 1,362,359 |  |  | * |  |  | <0.001 |
|  | Hispanic or Latino | 26,609 | 1.95 |  | * | 1.38 |  |  |
|  | Non-Hispanic Asian | 16,668 | 1.22 |  | * | 0.69 |  |  |
|  | Non-Hispanic Black or African American | 25,115 | 1.84 |  | * | 1.20 |  |  |
|  | Non-Hispanic White | 1,228,264 | 90.16 |  | * | 95.35 |  |  |
|  | Other including multiracial | 65,703 | 4.82 |  | * | 1.38 |  |  |
| Maryland | Total | 6,177,224 |  |  | 1,839 |  |  | <0.001 |
|  | Hispanic or Latino | 729,745 | 11.81 |  | 69 | 3.75 |  |  |
|  | Non-Hispanic Asian | 417,962 | 6.77 |  | 154 | 8.37 |  |  |
|  | Non-Hispanic Black or African American | 1,795,027 | 29.06 |  | 169 | 9.19 |  |  |
|  | Non-Hispanic White | 2,913,782 | 47.17 |  | 1,382 | 75.15 |  |  |
|  | Other including multiracial | 320,708 | 5.19 |  | 65 | 3.53 |  |  |
| Massachusetts | Total | 7,029,917 |  |  | 30,443 |  |  | <0.001 |
|  | Hispanic or Latino | 887,685 | 12.63 |  | 3,814 | 12.53 |  |  |
|  | Non-Hispanic Asian | 504,900 | 7.18 |  | 1,079 | 3.54 |  |  |
|  | Non-Hispanic Black or African American | 457,055 | 6.50 |  | 3,678 | 12.08 |  |  |
|  | Non-Hispanic White | 4,748,897 | 67.55 |  | 20,979 | 68.91 |  |  |
|  | Other including multiracial | 431,380 | 6.14 |  | 893 | 2.93 |  |  |
| Michigan | Total | 10,077,331 |  |  | 17,815 |  |  | <0.001 |
|  | Hispanic or Latino | 564,422 | 5.60 |  | 417 | 2.34 |  |  |
|  | Non-Hispanic Asian | 332,288 | 3.30 |  | 342 | 1.92 |  |  |
|  | Non-Hispanic Black or African American | 1,358,458 | 13.48 |  | 3,673 | 20.62 |  |  |
|  | Non-Hispanic White | 7,295,651 | 72.40 |  | 12,877 | 72.28 |  |  |
|  | Other including multiracial | 526,512 | 5.22 |  | 506 | 2.84 |  |  |
| Minnesota | Total | 5,706,494 |  |  | 3,625 |  |  | <0.001 |
|  | Hispanic or Latino | 345,640 | 6.06 |  | 30 | 0.83 |  |  |
|  | Non-Hispanic Asian | 297,460 | 5.21 |  | 69 | 1.90 |  |  |
|  | Non-Hispanic Black or African American | 392,850 | 6.88 |  | 73 | 2.01 |  |  |
|  | Non-Hispanic White | 4,353,880 | 76.30 |  | 3,408 | 94.01 |  |  |
|  | Other including multiracial | 316,664 | 5.55 |  | 45 | 1.24 |  |  |
| Mississippi | Total | 2,961,279 |  |  | * |  |  | <0.001 |
|  | Hispanic or Latino | 105,220 | 3.55 |  | * | 0.59 |  |  |
|  | Non-Hispanic Asian | 32,305 | 1.09 |  | * | 0.39 |  |  |
|  | Non-Hispanic Black or African American | 1,079,001 | 36.44 |  | * | 79.37 |  |  |
|  | Non-Hispanic White | 1,639,077 | 55.35 |  | * | 18.85 |  |  |
|  | Other including multiracial | 105,676 | 3.57 |  | * | 0.81 |  |  |
| Missouri | Total | 6,154,913 |  |  | * |  |  | <0.001 |
|  | Hispanic or Latino | 303,068 | 4.92 |  | * | 1.39 |  |  |
|  | Non-Hispanic Asian | 132,158 | 2.15 |  | * | 2.17 |  |  |
|  | Non-Hispanic Black or African American | 692,774 | 11.26 |  | * | 5.80 |  |  |
|  | Non-Hispanic White | 4,663,907 | 75.78 |  | * | 88.70 |  |  |
|  | Other including multiracial | 363,006 | 5.90 |  | * | 1.93 |  |  |
| Montana | Total | 1,084,225 |  |  | * |  |  | <0.001 |
|  | Hispanic or Latino | 45,199 | 4.17 |  | * | 0.78 |  |  |
|  | Non-Hispanic Asian | 8,077 | 0.74 |  | * | 0.39 |  |  |
|  | Non-Hispanic Black or African American | 5,077 | 0.47 |  | * | 1.56 |  |  |
|  | Non-Hispanic White | 901,318 | 83.13 |  | * | 96.89 |  |  |
|  | Other including multiracial | 124,554 | 11.49 |  | * | 0.39 |  |  |
| Nebraska | Total | 1,961,504 |  |  | * |  |  | <0.001 |
|  | Hispanic or Latino | 234,715 | 11.97 |  | * | 29.28 |  |  |
|  | Non-Hispanic Asian | 52,359 | 2.67 |  | * | 1.87 |  |  |
|  | Non-Hispanic Black or African American | 94,405 | 4.81 |  | * | 3.12 |  |  |
|  | Non-Hispanic White | 1,484,687 | 75.69 |  | * | 63.86 |  |  |
|  | Other including multiracial | 95,338 | 4.86 |  | * | 1.87 |  |  |
| Nevada | Total | 3,104,614 |  |  | 570 |  |  | <0.001 |
|  | Hispanic or Latino | 890,257 | 28.68 |  | 55 | 9.65 |  |  |
|  | Non-Hispanic Asian | 265,991 | 8.57 |  | 26 | 4.56 |  |  |
|  | Non-Hispanic Black or African American | 291,960 | 9.40 |  | 39 | 6.84 |  |  |
|  | Non-Hispanic White | 1,425,952 | 45.93 |  | 423 | 74.21 |  |  |
|  | Other including multiracial | 230,454 | 7.42 |  | 27 | 4.74 |  |  |
| New Hampshire | Total | 1,377,529 |  |  | * |  |  | <0.001 |
|  | Hispanic or Latino | 59,454 | 4.32 |  | * | 2.81 |  |  |
|  | Non-Hispanic Asian | 35,604 | 2.58 |  | * | 0.86 |  |  |
|  | Non-Hispanic Black or African American | 18,655 | 1.35 |  | * | 0.43 |  |  |
|  | Non-Hispanic White | 1,200,649 | 87.16 |  | * | 94.28 |  |  |
|  | Other including multiracial | 63,167 | 4.59 |  | * | 1.62 |  |  |
| New Jersey | Total | 9,288,994 |  |  | 2,040 |  |  | <0.001 |
|  | Hispanic or Latino | 2,002,575 | 21.56 |  | 283 | 13.87 |  |  |
|  | Non-Hispanic Asian | 942,921 | 10.15 |  | 239 | 11.72 |  |  |
|  | Non-Hispanic Black or African American | 1,154,142 | 12.42 |  | 147 | 7.21 |  |  |
|  | Non-Hispanic White | 4,816,381 | 51.85 |  | 1,299 | 63.68 |  |  |
|  | Other including multiracial | 372,975 | 4.02 |  | 72 | 3.53 |  |  |
| New Mexico | Total | 2,117,522 |  |  | * |  |  | <0.001 |
|  | Hispanic or Latino | 1,010,811 | 47.74 |  | * | 22.81 |  |  |
|  | Non-Hispanic Asian | 35,261 | 1.67 |  | * | 1.63 |  |  |
|  | Non-Hispanic Black or African American | 38,330 | 1.81 |  | * | 3.85 |  |  |
|  | Non-Hispanic White | 772,952 | 36.50 |  | * | 69.04 |  |  |
|  | Other including multiracial | 260,168 | 12.29 |  | * | 2.67 |  |  |
| New York | Total | 20,201,249 |  |  | 30,017 |  |  | <0.001 |
|  | Hispanic or Latino | 3,948,032 | 19.54 |  | 11,429 | 38.08 |  |  |
|  | Non-Hispanic Asian | 1,916,329 | 9.49 |  | 1,536 | 5.12 |  |  |
|  | Non-Hispanic Black or African American | 2,759,022 | 13.66 |  | 6,186 | 20.61 |  |  |
|  | Non-Hispanic White | 10,598,907 | 52.47 |  | 10,013 | 33.36 |  |  |
|  | Other including multiracial | 978,959 | 4.85 |  | 853 | 2.84 |  |  |
| North Carolina | Total | 10,439,388 |  |  | 1,504 |  |  | <0.001 |
|  | Hispanic or Latino | 1,118,596 | 10.72 |  | 96 | 6.38 |  |  |
|  | Non-Hispanic Asian | 340,059 | 3.26 |  | 56 | 3.72 |  |  |
|  | Non-Hispanic Black or African American | 2,107,526 | 20.19 |  | 162 | 10.77 |  |  |
|  | Non-Hispanic White | 6,312,148 | 60.46 |  | 1,155 | 76.80 |  |  |
|  | Other including multiracial | 561,059 | 5.37 |  | 35 | 2.33 |  |  |
| North Dakota | Total | 779,094 |  |  | * |  |  | <0.001 |
|  | Hispanic or Latino | 33,412 | 4.29 |  | * | 2.93 |  |  |
|  | Non-Hispanic Asian | 13,050 | 1.68 |  | * | 0.98 |  |  |
|  | Non-Hispanic Black or African American | 26,152 | 3.36 |  | * | 1.95 |  |  |
|  | Non-Hispanic White | 636,160 | 81.65 |  | * | 92.83 |  |  |
|  | Other including multiracial | 70,320 | 9.03 |  | * | 1.30 |  |  |
| Ohio | Total | 11,799,448 |  |  | 1,266 |  |  | <0.001 |
|  | Hispanic or Latino | 521,308 | 4.42 |  | 23 | 1.82 |  |  |
|  | Non-Hispanic Asian | 296,604 | 2.51 |  | 44 | 3.48 |  |  |
|  | Non-Hispanic Black or African American | 1,457,180 | 12.35 |  | 73 | 5.77 |  |  |
|  | Non-Hispanic White | 8,954,135 | 75.89 |  | 1,100 | 86.89 |  |  |
|  | Other including multiracial | 570,221 | 4.83 |  | 26 | 2.05 |  |  |
| Oklahoma | Total | 3,959,353 |  |  | * |  |  | <0.001 |
|  | Hispanic or Latino | 471,931 | 11.92 |  | * | 2.61 |  |  |
|  | Non-Hispanic Asian | 89,653 | 2.26 |  | * | 2.94 |  |  |
|  | Non-Hispanic Black or African American | 283,242 | 7.15 |  | * | 7.84 |  |  |
|  | Non-Hispanic White | 2,407,188 | 60.80 |  | * | 83.66 |  |  |
|  | Other including multiracial | 707,339 | 17.87 |  | * | 2.94 |  |  |
| Oregon | Total | 4,237,256 |  |  | * |  |  | <0.001 |
|  | Hispanic or Latino | 588,757 | 13.89 |  | * | 2.04 |  |  |
|  | Non-Hispanic Asian | 191,797 | 4.53 |  | * | 4.08 |  |  |
|  | Non-Hispanic Black or African American | 78,658 | 1.86 |  | * | 0.91 |  |  |
|  | Non-Hispanic White | 3,036,158 | 71.65 |  | * | 89.69 |  |  |
|  | Other including multiracial | 341,886 | 8.07 |  | * | 3.28 |  |  |
| Pennsylvania | Total | 13,002,700 |  |  | 32,709 |  |  | <0.001 |
|  | Hispanic or Latino | 1,049,615 | 8.07 |  | 304 | 0.93 |  |  |
|  | Non-Hispanic Asian | 506,674 | 3.90 |  | 828 | 2.53 |  |  |
|  | Non-Hispanic Black or African American | 1,368,978 | 10.53 |  | 4,169 | 12.75 |  |  |
|  | Non-Hispanic White | 9,553,417 | 73.47 |  | 26,767 | 81.83 |  |  |
|  | Other including multiracial | 524,016 | 4.03 |  | 641 | 1.96 |  |  |
| Puerto Rico | Total | 3,285,874 |  |  | * |  |  | <0.001 |
|  | Hispanic or Latino | 3,249,043 | 98.88 |  | * | 68.18 |  |  |
|  | Non-Hispanic Asian | 2,746 | 0.08 |  | * | 1.14 |  |  |
|  | Non-Hispanic Black or African American | 4,286 | 0.13 |  | * | 1.14 |  |  |
|  | Non-Hispanic White | 24,548 | 0.75 |  | * | 28.41 |  |  |
|  | Other including multiracial | 5,251 | 0.16 |  | * | 1.14 |  |  |
| Rhode Island | Total | 1,097,379 |  |  | * |  |  | <0.001 |
|  | Hispanic or Latino | 182,101 | 16.59 |  | * | 3.07 |  |  |
|  | Non-Hispanic Asian | 38,367 | 3.50 |  | * | 1.23 |  |  |
|  | Non-Hispanic Black or African American | 55,386 | 5.05 |  | * | 3.48 |  |  |
|  | Non-Hispanic White | 754,050 | 68.71 |  | * | 89.96 |  |  |
|  | Other including multiracial | 67,475 | 6.15 |  | * | 2.25 |  |  |
| South Carolina | Total | 5,118,425 |  |  | * |  |  | <0.001 |
|  | Hispanic or Latino | 352,838 | 6.89 |  | * | 17.47 |  |  |
|  | Non-Hispanic Asian | 89,394 | 1.75 |  | * | 0.35 |  |  |
|  | Non-Hispanic Black or African American | 1,269,031 | 24.79 |  | * | 50.77 |  |  |
|  | Non-Hispanic White | 3,178,552 | 62.10 |  | * | 30.52 |  |  |
|  | Other including multiracial | 228,610 | 4.47 |  | * | 0.88 |  |  |
| South Dakota | Total | 886,667 |  |  | * |  |  | <0.001 |
|  | Hispanic or Latino | 38,741 | 4.37 |  | * | 0.50 |  |  |
|  | Non-Hispanic Asian | 13,332 | 1.50 |  | * | 1.49 |  |  |
|  | Non-Hispanic Black or African American | 17,441 | 1.97 |  | * | 1.98 |  |  |
|  | Non-Hispanic White | 705,583 | 79.58 |  | * | 93.56 |  |  |
|  | Other including multiracial | 111,570 | 12.58 |  | * | 2.48 |  |  |
| Tennessee | Total | 6,910,840 |  |  | 2,567 |  |  | <0.001 |
|  | Hispanic or Latino | 479,187 | 6.93 |  | 111 | 4.32 |  |  |
|  | Non-Hispanic Asian | 134,302 | 1.94 |  | 44 | 1.71 |  |  |
|  | Non-Hispanic Black or African American | 1,083,772 | 15.68 |  | 413 | 16.09 |  |  |
|  | Non-Hispanic White | 4,900,246 | 70.91 |  | 1,955 | 76.16 |  |  |
|  | Other including multiracial | 313,333 | 4.53 |  | 44 | 1.71 |  |  |
| Texas | Total | 29,145,505 |  |  | 11,889 |  |  | <0.001 |
|  | Hispanic or Latino | 11,441,717 | 39.26 |  | 1,762 | 14.82 |  |  |
|  | Non-Hispanic Asian | 1,561,518 | 5.36 |  | 410 | 3.45 |  |  |
|  | Non-Hispanic Black or African American | 3,444,712 | 11.82 |  | 2,198 | 18.49 |  |  |
|  | Non-Hispanic White | 11,584,597 | 39.75 |  | 7,321 | 61.58 |  |  |
|  | Other including multiracial | 1,112,961 | 3.82 |  | 198 | 1.67 |  |  |
| Utah | Total | 3,271,616 |  |  | * |  |  | <0.001 |
|  | Hispanic or Latino | 492,912 | 15.07 |  | * | 8.32 |  |  |
|  | Non-Hispanic Asian | 78,618 | 2.40 |  | * | 1.39 |  |  |
|  | Non-Hispanic Black or African American | 37,192 | 1.14 |  | * | 0.99 |  |  |
|  | Non-Hispanic White | 2,465,355 | 75.36 |  | * | 86.34 |  |  |
|  | Other including multiracial | 197,539 | 6.04 |  | * | 2.97 |  |  |
| Vermont | Total | 643,077 |  |  | * |  |  | 0.004 |
|  | Hispanic or Latino | 15,504 | 2.41 |  | * | 0.00 |  |  |
|  | Non-Hispanic Asian | 11,457 | 1.78 |  | * | 0.53 |  |  |
|  | Non-Hispanic Black or African American | 8,649 | 1.34 |  | * | 0.00 |  |  |
|  | Non-Hispanic White | 573,201 | 89.13 |  | * | 97.86 |  |  |
|  | Other including multiracial | 34,266 | 5.33 |  | * | 1.60 |  |  |
| Virginia | Total | 8,631,393 |  |  | 1,538 |  |  | <0.001 |
|  | Hispanic or Latino | 908,749 | 10.53 |  | 54 | 3.51 |  |  |
|  | Non-Hispanic Asian | 610,612 | 7.07 |  | 104 | 6.76 |  |  |
|  | Non-Hispanic Black or African American | 1,578,090 | 18.28 |  | 106 | 6.89 |  |  |
|  | Non-Hispanic White | 5,058,363 | 58.60 |  | 1,229 | 79.91 |  |  |
|  | Other including multiracial | 475,579 | 5.51 |  | 45 | 2.93 |  |  |
| Washington | Total | 7,705,281 |  |  | 2,057 |  |  | <0.001 |
|  | Hispanic or Latino | 1,059,213 | 13.75 |  | 70 | 3.40 |  |  |
|  | Non-Hispanic Asian | 723,062 | 9.38 |  | 75 | 3.65 |  |  |
|  | Non-Hispanic Black or African American | 296,170 | 3.84 |  | 25 | 1.22 |  |  |
|  | Non-Hispanic White | 4,918,820 | 63.84 |  | 1,843 | 89.60 |  |  |
|  | Other including multiracial | 708,016 | 9.19 |  | 44 | 2.14 |  |  |
| West Virginia | Total | 1,793,716 |  |  | * |  |  | 0.226 |
|  | Hispanic or Latino | 34,827 | 1.94 |  | * | 1.16 |  |  |
|  | Non-Hispanic Asian | 14,903 | 0.83 |  | * | 0.58 |  |  |
|  | Non-Hispanic Black or African American | 64,749 | 3.61 |  | * | 4.62 |  |  |
|  | Non-Hispanic White | 1,598,834 | 89.14 |  | * | 92.49 |  |  |
|  | Other including multiracial | 80,403 | 4.48 |  | * | 1.16 |  |  |
| Wisconsin | Total | 5,893,718 |  |  | 20,318 |  |  | <0.001 |
|  | Hispanic or Latino | 447,290 | 7.59 |  | 808 | 3.98 |  |  |
|  | Non-Hispanic Asian | 174,267 | 2.96 |  | 361 | 1.78 |  |  |
|  | Non-Hispanic Black or African American | 366,508 | 6.22 |  | 1,540 | 7.58 |  |  |
|  | Non-Hispanic White | 4,634,018 | 78.63 |  | 17,363 | 85.46 |  |  |
|  | Other including multiracial | 271,635 | 4.61 |  | 246 | 1.21 |  |  |
| Wyoming | Total | 576,851 |  |  | * |  |  | <0.001 |
|  | Hispanic or Latino | 59,046 | 10.24 |  | * | 1.28 |  |  |
|  | Non-Hispanic Asian | 5,037 | 0.87 |  | * | 1.28 |  |  |
|  | Non-Hispanic Black or African American | 4,735 | 0.82 |  | * | 3.85 |  |  |
|  | Non-Hispanic White | 469,664 | 81.42 |  | * | 93.59 |  |  |
|  | Other including multiracial | 38,369 | 6.65 |  | * | 0.00 |  |  |
| ^a^Per *All of Us* Data and Statistics Dissemination Policy, participant count of 1 to 20 nor data/statistics that allow a participant count of 1 to 20 to be derived from other reported information cannot be published directly—thus indicated as *. | | | | | | | | |
| ^b^*P* value derived from Chi-square difference test | |  |  |  |  |  |  |  |

| Table S2. Differences in Hispanic or Latino representation between the US and *All of Us* | | | | | | | |  |
| --- | --- | --- | --- | --- | --- | --- | --- | --- |
|  | US | |  | *All of Us* | |  | Difference (95% CI), %^b^ |  |
| State | Count | Proportion, % | | Count | Proportion, % | | (*All of Us* - US) | *P* value^c^ |
| Alabama | 264,047 | 5.26 |  | 382 | 1.77 |  | -3.49 (-3.67, -3.31) | <0.001 |
| Alaska | 49,824 | 6.79 |  | * | 5.63 |  | -1.17 (-4.74, 2.40) | 0.557 |
| Arizona | 2,192,253 | 30.65 |  | 13,486 | 30.34 |  | -0.31 (-0.07, 0.13) | 0.175 |
| Arkansas | 256,847 | 8.53 |  | 38 | 9.09 |  | 0.56 (-2.19, 3.32) | 0.681 |
| California | 15,579,652 | 39.40 |  | 13,400 | 28.01 |  | -11.40 (-11.80, -10.99) | <0.001 |
| Colorado | 1,263,390 | 21.88 |  | 220 | 13.74 |  | -8.14 (-9.83, -6.46) | <0.001 |
| Connecticut | 623,293 | 17.29 |  | 791 | 32.10 |  | 14.82 (12.97, 16.61) | <0.001 |
| Delaware | 104,290 | 10.53 |  | * | 8.57 |  | -1.96 (-6.60, 2.67) | 0.449 |
| District of Columbia | 77,652 | 11.26 |  | 23 | 5.26 |  | -6.00 (-8.09, -3.90) | <0.001 |
| Florida | 5,697,240 | 26.45 |  | 5,697 | 33.27 |  | 6.82 (6.11, 7.53) | <0.001 |
| Georgia | 1,123,457 | 10.49 |  | 165 | 1.83 |  | -8.66 (-8.94, -8.38) | <0.001 |
| Hawaii | 138,923 | 9.55 |  | * | 4.35 |  | -5.20 (-8.15, -2.25) | <0.001 |
| Idaho | 239,407 | 13.02 |  | * | 4.08 |  | -8.94 (-11.41, -6.46) | <0.001 |
| Illinois | 2,337,410 | 18.24 |  | 3,882 | 11.89 |  | -6.35 (-6.70, -6.00) | <0.001 |
| Indiana | 554,191 | 8.17 |  | 42 | 3.62 |  | -4.55 (-5.62, -3.48) | <0.001 |
| Iowa | 215,986 | 6.77 |  | * | 0.94 |  | -5.83 (-6.65, -5.01) | <0.001 |
| Kansas | 382,603 | 13.02 |  | 27 | 2.74 |  | -10.28 (-11.30, -9.26) | <0.001 |
| Kentucky | 207,854 | 4.61 |  | * | 1.53 |  | -3.08 (-4.30, -1.86) | 0.004 |
| Louisiana | 322,549 | 6.92 |  | 84 | 2.16 |  | -4.76 (-5.22, -4.30) | <0.001 |
| Maine | 26,609 | 1.95 |  | * | 1.38 |  | -0.58 (-1.52, 0.37) | 0.316 |
| Maryland | 729,745 | 11.81 |  | 69 | 3.75 |  | -8.06 (-8.93, -7.19) | <0.001 |
| Massachusetts | 887,685 | 12.63 |  | 3,814 | 12.53 |  | -0.10 (-0.47, 0.27) | 0.604 |
| Michigan | 564,422 | 5.60 |  | 417 | 2.34 |  | -3.26 (-3.48, -3.04) | <0.001 |
| Minnesota | 345,640 | 6.06 |  | 30 | 0.83 |  | -5.23 (-5.52, -4.93) | <0.001 |
| Mississippi | 105,220 | 3.55 |  | 21 | 0.59 |  | -2.97 (-3.22, -2.72) | <0.001 |
| Missouri | 303,068 | 4.92 |  | * | 1.39 |  | -3.53 (-4.17, -2.89) | <0.001 |
| Montana | 45,199 | 4.17 |  | * | 0.78 |  | -3.59 (-4.46, -2.32) | 0.007 |
| Nebraska | 234,715 | 11.97 |  | 94 | 29.28 |  | 17.32 (12.34, 22.30) | <0.001 |
| Nevada | 890,257 | 28.68 |  | 55 | 9.65 |  | -19.03 (-21.45, -16.60) | <0.001 |
| New Hampshire | 59,454 | 4.32 |  | 26 | 2.81 |  | -1.51 (-2.57, -0.44) | 0.024 |
| New Jersey | 2,002,575 | 21.56 |  | 283 | 13.87 |  | -7.69 (-9.19, -6.19) | <0.001 |
| New Mexico | 1,010,811 | 47.74 |  | 154 | 22.81 |  | -24.92 (-28.09, -21.75) | <0.001 |
| New York | 3,948,032 | 19.54 |  | 11,429 | 38.08 |  | 18.53 (17.98, 19.08) | <0.001 |
| North Carolina | 1,118,596 | 10.72 |  | 96 | 6.38 |  | -4.33 (-5.57, -3.10) | <0.001 |
| North Dakota | 33,412 | 4.29 |  | * | 2.93 |  | -1.36 (-3.24, 0.53) | 0.241 |
| Ohio | 521,308 | 4.42 |  | 23 | 1.82 |  | -2.60 (-3.34, -1.87) | <0.001 |
| Oklahoma | 471,931 | 11.92 |  | * | 2.61 |  | -9.31 (-11.09, -7.52) | <0.001 |
| Oregon | 588,757 | 13.89 |  | * | 2.04 |  | -11.86 (-12.79, -10.92) | <0.001 |
| Pennsylvania | 1,049,615 | 8.07 |  | 304 | 0.93 |  | -7.14 (-7.25, -7.04) | <0.001 |
| Puerto Rico | 3,249,043 | 98.88 |  | 60 | 68.18 |  | -30.70 (-40.43, -20.97) | <0.001 |
| Rhode Island | 182,101 | 16.59 |  | * | 3.07 |  | -13.52 (-15.05, -11.99) | <0.001 |
| South Carolina | 352,838 | 6.89 |  | 396 | 17.47 |  | 10.57 (9.01, 12.14) | <0.001 |
| South Dakota | 38,741 | 4.37 |  | * | 0.50 |  | -3.87 (-4.84, -2.91) | 0.007 |
| Tennessee | 479,187 | 6.93 |  | 111 | 4.32 |  | -2.61 (-3.40, -1.82) | <0.001 |
| Texas | 11,441,717 | 39.26 |  | 1,762 | 14.82 |  | -24.44 (-25.08, -23.80) | <0.001 |
| Utah | 492,912 | 15.07 |  | 42 | 8.32 |  | -6.75 (-9.16, -4.34) | <0.001 |
| Vermont | 15,504 | 2.41 |  | 0 | 0.00 |  | -2.41 (-2.44, -2.37) | <0.001 |
| Virginia | 908,749 | 10.53 |  | 54 | 3.51 |  | -7.02 (-7.94, -6.10) | <0.001 |
| Washington | 1,059,213 | 13.75 |  | 70 | 3.40 |  | -10.34 (-11.12, -9.56) | <0.001 |
| West Virginia | 34,827 | 1.94 |  | * | 1.16 |  | -0.79 (-2.38, 0.81) | 0.454 |
| Wisconsin | 447,290 | 7.59 |  | 808 | 3.98 |  | -3.61 (-3.88, -3.34) | <0.001 |
| Wyoming | 59,046 | 10.24 |  | * | 1.28 |  | -8.95 (-10.72, -7.19) | <0.001 |
| ^a^Per *All of Us* Data and Statistics Dissemination Policy, a participant count of 1 to 20 nor data/statistics that allow a participant count of 1 to 20 to be derived from other reported information cannot be published directly—thus indicated as *. | | | | | | | | |
| ^b^Positive value indicates overrepresentation in *All of Us*; negative value indicates underrepresentation in *All of Us*. | | | | | | | | |
| ^c^Calculated using the 2-sample test for equality of proportions | | | | |  |  |  |  |

| Table S3. Differences in non-Hispanic Asian representation between the US and *All of Us* | | | | | | | |  |
| --- | --- | --- | --- | --- | --- | --- | --- | --- |
|  | US | |  | *All of Us* | |  | Difference (95% CI), %^b^ |  |
| State | Count | Proportion, % | | Count | Proportion, % | | (*All of Us* - US) | *P* value^c^ |
| Alabama | 75,918 | 1.51 |  | 236 | 1.09 |  | -0.42 (-0.56, -0.28) | <0.001 |
| Alaska | 43,449 | 5.92 |  | * | 3.13 |  | -2.80 (-5.50, -1.03) | 0.134 |
| Arizona | 248,837 | 3.48 |  | 690 | 1.55 |  | -1.93 (-2.04, -1.81) | <0.001 |
| Arkansas | 51,210 | 1.70 |  | * | 1.91 |  | 0.21 (-1.10, 1.53) | 0.736 |
| California | 5,978,795 | 15.12 |  | 4,553 | 9.52 |  | -5.60 (-5.86, -5.34) | <0.001 |
| Colorado | 195,220 | 3.38 |  | 35 | 2.19 |  | -1.20 (-1.91, -0.48) | 0.008 |
| Connecticut | 170,459 | 4.73 |  | 31 | 1.26 |  | -3.47 (-3.91, -3.03) | <0.001 |
| Delaware | 42,398 | 4.28 |  | * | 2.86 |  | -1.43 (-4.19, 1.33) | 0.405 |
| District of Columbia | 33,192 | 4.81 |  | 23 | 5.26 |  | 0.45 (-1.64, 2.54) | 0.661 |
| Florida | 629,626 | 2.92 |  | 339 | 1.98 |  | -0.94 (-1.15, -0.73) | <0.001 |
| Georgia | 475,680 | 4.44 |  | 289 | 3.20 |  | -1.24 (-1.60, -0.87) | <0.001 |
| Hawaii | 531,558 | 36.53 |  | 30 | 16.30 |  | -20.22 (-25.56, -14.88) | <0.001 |
| Idaho | 26,036 | 1.42 |  | * | 0.41 |  | -1.01 (-1.81, -0.21) | 0.182 |
| Illinois | 747,280 | 5.83 |  | 815 | 2.50 |  | -3.34 (-3.50, -3.17) | <0.001 |
| Indiana | 166,651 | 2.46 |  | 27 | 2.33 |  | -0.13 (-1.00, 0.74) | 0.774 |
| Iowa | 75,017 | 2.35 |  | * | 1.69 |  | -0.66 (-1.76, 0.44) | 0.315 |
| Kansas | 85,225 | 2.90 |  | * | 0.81 |  | -2.09 (-2.65, -1.53) | <0.001 |
| Kentucky | 73,843 | 1.64 |  | * | 1.79 |  | 0.15 (-1.16, 1.47) | 0.814 |
| Louisiana | 85,336 | 1.83 |  | 84 | 2.16 |  | 0.33 (-0.13, 0.79) | 0.125 |
| Maine | 16,668 | 1.22 |  | * | 0.69 |  | -0.53 (-1.21, 0.14) | 0.241 |
| Maryland | 417,962 | 6.77 |  | 154 | 8.37 |  | 1.61 (0.34, 2.87) | 0.006 |
| Massachusetts | 504,900 | 7.18 |  | 1,079 | 3.54 |  | -3.64 (-3.85, -3.43) | <0.001 |
| Michigan | 332,288 | 3.30 |  | 342 | 1.92 |  | -1.38 (-1.58, -1.18) | <0.001 |
| Minnesota | 297,460 | 5.21 |  | 69 | 1.90 |  | -3.31 (-3.75, -2.86) | <0.001 |
| Mississippi | 32,305 | 1.09 |  | * | 0.39 |  | -0.70 (-0.91, -0.50) | <0.001 |
| Missouri | 132,158 | 2.15 |  | 28 | 2.17 |  | 0.02 (-0.77, 0.81) | 0.961 |
| Montana | 8,077 | 0.74 |  | * | 0.39 |  | -0.36 (-1.12, 0.41) | 0.507 |
| Nebraska | 52,359 | 2.67 |  | * | 1.87 |  | -0.80 (-2.28, 0.68) | 0.374 |
| Nevada | 265,991 | 8.57 |  | 26 | 4.56 |  | -4.01 (-5.72, -2.29) | 0.001 |
| New Hampshire | 35,604 | 2.58 |  | * | 0.86 |  | -1.72 (-2.32, -1.12) | 0.001 |
| New Jersey | 942,921 | 10.15 |  | 239 | 11.72 |  | 1.56 (0.17, 2.96) | 0.019 |
| New Mexico | 35,261 | 1.67 |  | * | 1.63 |  | -0.04 (-0.99, 0.92) | 0.942 |
| New York | 1,916,329 | 9.49 |  | 1,536 | 5.12 |  | -4.37 (-4.62, -4.12) | <0.001 |
| North Carolina | 340,059 | 3.26 |  | 56 | 3.72 |  | 0.47 (-0.49, 1.42) | 0.309 |
| North Dakota | 13,050 | 1.68 |  | * | 0.98 |  | -0.70 (-1.80, 0.40) | 0.341 |
| Ohio | 296,604 | 2.51 |  | 44 | 3.48 |  | 0.96 (-0.05, 1.97) | 0.029 |
| Oklahoma | 89,653 | 2.26 |  | * | 2.94 |  | 0.68 (-1.22, 2.57) | 0.426 |
| Oregon | 191,797 | 4.53 |  | 36 | 4.08 |  | -0.45 (-1.75, 0.86) | 0.521 |
| Pennsylvania | 506,674 | 3.90 |  | 828 | 2.53 |  | -1.37 (-1.54, -1.19) | <0.001 |
| Puerto Rico | 2,746 | 0.08 |  | * | 1.14 |  | 1.05 (-1.16, 3.27) | 0.001 |
| Rhode Island | 38,367 | 3.50 |  | * | 1.23 |  | -2.27 (-3.25, -1.29) | 0.006 |
| South Carolina | 89,394 | 1.75 |  | * | 0.35 |  | -1.39 (-1.64, -1.15) | <0.001 |
| South Dakota | 13,332 | 1.50 |  | * | 1.49 |  | -0.02 (-1.69, 1.65) | 0.983 |
| Tennessee | 134,302 | 1.94 |  | 44 | 1.71 |  | -0.23 (-0.73, 0.27) | 0.400 |
| Texas | 1,561,518 | 5.36 |  | 410 | 3.45 |  | -1.91 (-2.24, -1.58) | <0.001 |
| Utah | 78,618 | 2.40 |  | * | 1.39 |  | -1.02 (-2.04, 0.00) | 0.136 |
| Vermont | 11,457 | 1.78 |  | * | 0.53 |  | -1.25 (-2.29, -0.20) | 0.197 |
| Virginia | 610,612 | 7.07 |  | 104 | 6.76 |  | -0.31 (-1.57, 0.94) | 0.633 |
| Washington | 723,062 | 9.38 |  | 75 | 3.65 |  | -5.74 (-6.55, -4.93) | <0.001 |
| West Virginia | 14,903 | 0.83 |  | * | 0.58 |  | -0.25 (-1.38, 0.88) | 0.714 |
| Wisconsin | 174,267 | 2.96 |  | 361 | 1.78 |  | -1.18 (-1.36, -1.00) | <0.001 |
| Wyoming | 5,037 | 0.87 |  | * | 1.28 |  | 0.41 (-1.36, 2.17) | 0.583 |
| ^a^Per *All of Us* Data and Statistics Dissemination Policy, a participant count of 1 to 20 nor data/statistics that allow a participant count of 1 to 20 to be derived from other reported information cannot be published directly—thus indicated as *. | | | | | | | | |
| ^b^Positive value indicates overrepresentation in *All of Us*; negative value indicates underrepresentation in *All of Us*. | | | | | | | | |
| ^c^Calculated using the 2-sample test for equality of proportions | | | | |  |  |  |  |

| Table S4. Differences in non-Hispanic Black or African American representation between the US and *All of Us* | | | | | | | | |
| --- | --- | --- | --- | --- | --- | --- | --- | --- |
|  | US | |  | *All of Us* | |  | Difference (95% CI), %^b^ |  |
| State | Count | Proportion, % | | Count | Proportion, % | | (*All of Us* - US) | *P* value^c^ |
| Alabama | 1,288,159 | 25.64 |  | 11,926 | 55.11 |  | 29.47 (28.81, 30.14) | <0.001 |
| Alaska | 20,731 | 2.83 |  | 27 | 16.88 |  | 14.05 (8.24, 19.85) | <0.001 |
| Arizona | 317,161 | 4.43 |  | 3,865 | 8.70 |  | 4.26 (4.00, 4.52) | <0.001 |
| Arkansas | 449,884 | 14.94 |  | 91 | 21.77 |  | 6.83 (2.88, 10.79) | <0.001 |
| California | 2,119,286 | 5.36 |  | 2,289 | 4.78 |  | -0.58 (-0.77, -0.38) | <0.001 |
| Colorado | 221,310 | 3.83 |  | 38 | 2.37 |  | -1.46 (-2.21, -0.71) | <0.001 |
| Connecticut | 360,937 | 10.01 |  | 296 | 12.01 |  | 2.00 (0.72, 3.29) | 0.001 |
| Delaware | 212,960 | 21.51 |  | * | 7.86 |  | -13.66 (-18.11, -9.20) | <0.001 |
| District of Columbia | 282,066 | 40.91 |  | 41 | 9.38 |  | -31.52 (-34.26, -28.79) | <0.001 |
| Florida | 3,127,052 | 14.52 |  | 4,297 | 25.09 |  | 10.58 (9.93, 11.23) | <0.001 |
| Georgia | 3,278,119 | 30.60 |  | 4,557 | 50.53 |  | 19.93 (18.90, 20.96) | <0.001 |
| Hawaii | 21,877 | 1.50 |  | * | 1.63 |  | 0.13 (-1.70, 1.96) | 0.887 |
| Idaho | 14,785 | 0.80 |  | * | 2.04 |  | 1.24 (-0.53, 3.01) | 0.030 |
| Illinois | 1,775,612 | 13.86 |  | 16,357 | 50.12 |  | 36.26 (35.72, 36.80) | <0.001 |
| Indiana | 637,500 | 9.39 |  | 147 | 12.66 |  | 3.27 (1.36, 5.18) | <0.001 |
| Iowa | 129,321 | 4.05 |  | * | 3.20 |  | -0.86 (-2.35, 0.64) | 0.316 |
| Kansas | 163,352 | 5.56 |  | 82 | 8.32 |  | 2.76 (1.04, 4.49) | <0.001 |
| Kentucky | 357,764 | 7.94 |  | 20 | 5.12 |  | -2.82 (-5.01, -0.64) | 0.039 |
| Louisiana | 1,452,420 | 31.18 |  | 2,425 | 62.42 |  | 31.24 (29.71, 32.76) | <0.001 |
| Maine | 25,115 | 1.84 |  | * | 1.20 |  | -0.64 (-1.53, 0.25) | 0.253 |
| Maryland | 1,795,027 | 29.06 |  | 169 | 9.19 |  | -19.87 (-21.19, -18.55) | <0.001 |
| Massachusetts | 457,055 | 6.50 |  | 3,678 | 12.08 |  | 5.58 (5.21, 5.95) | <0.001 |
| Michigan | 1,358,458 | 13.48 |  | 3,673 | 20.62 |  | 7.14 (6.54, 7.73) | <0.001 |
| Minnesota | 392,850 | 6.88 |  | 73 | 2.01 |  | -4.87 (-5.33, -4.41) | <0.001 |
| Mississippi | 1,079,001 | 36.44 |  | 2,847 | 79.37 |  | 42.93 (41.61, 44.26) | <0.001 |
| Missouri | 692,774 | 11.26 |  | 75 | 5.80 |  | -5.45 (-6.73, -4.18) | <0.001 |
| Montana | 5,077 | 0.47 |  | * | 1.56 |  | 1.09 (-0.43, 2.60) | 0.011 |
| Nebraska | 94,405 | 4.81 |  | * | 3.12 |  | -1.70 (-3.60, 0.20) | 0.155 |
| Nevada | 291,960 | 9.40 |  | 39 | 6.84 |  | -2.56 (-4.63, -0.49) | 0.036 |
| New Hampshire | 18,655 | 1.35 |  | * | 0.43 |  | -0.92 (-1.35, -0.50) | <0.001 |
| New Jersey | 1,154,142 | 12.42 |  | 147 | 7.21 |  | -5.22 (-6.34, -4.10) | <0.001 |
| New Mexico | 38,330 | 1.81 |  | 26 | 3.85 |  | 2.04 (0.59, 3.49) | <0.001 |
| New York | 2,759,022 | 13.66 |  | 6,186 | 20.61 |  | 6.95 (6.59, 7.41) | <0.001 |
| North Carolina | 2,107,526 | 20.19 |  | 162 | 10.77 |  | -9.42 (-10.98, -7.85) | <0.001 |
| North Dakota | 26,152 | 3.36 |  | * | 1.95 |  | -1.40 (-2.95, 0.15) | 0.173 |
| Ohio | 1,457,180 | 12.35 |  | 73 | 5.77 |  | -6.58 (-7.87, -5.30) | <0.001 |
| Oklahoma | 283,242 | 7.15 |  | 24 | 7.84 |  | 0.69 (-2.32, 3.70) | 0.640 |
| Oregon | 78,658 | 1.86 |  | * | 0.91 |  | -0.95 (-1.58, -0.33) | 0.036 |
| Pennsylvania | 1,368,978 | 10.53 |  | 4,169 | 12.75 |  | 2.22 (1.86, 2.58) | <0.001 |
| Puerto Rico | 4,286 | 0.13 |  | * | 1.14 |  | 1.01 (-1.21, 3.22) | 0.009 |
| Rhode Island | 55,386 | 5.05 |  | * | 3.48 |  | -1.56 (-3.19, 0.06) | 0.115 |
| South Carolina | 1,269,031 | 24.79 |  | 1,151 | 50.77 |  | 25.98 (23.92, 28.04) | <0.001 |
| South Dakota | 17,441 | 1.97 |  | * | 1.98 |  | 0.01 (-1.91, 1.93) | 0.989 |
| Tennessee | 1,083,772 | 15.68 |  | 413 | 16.09 |  | 0.41 (-1.02, 1.83) | 0.571 |
| Texas | 3,444,712 | 11.82 |  | 2,198 | 18.49 |  | 6.67 (5.97, 7.37) | <0.001 |
| Utah | 37,192 | 1.14 |  | * | 0.99 |  | -0.15 (-1.01, 0.72) | 0.756 |
| Vermont | 8,649 | 1.34 |  | 0 | 0.00 |  | -1.34 (-1.37, -1.32) | 0.110 |
| Virginia | 1,578,090 | 18.28 |  | 106 | 6.89 |  | -11.39 (-12.66, -10.12) | <0.001 |
| Washington | 296,170 | 3.84 |  | 25 | 1.22 |  | -2.63 (-3.10, -2.15) | <0.001 |
| West Virginia | 64,749 | 3.61 |  | * | 4.62 |  | 1.01 (-2.12, 4.14) | 0.474 |
| Wisconsin | 366,508 | 6.22 |  | 1,540 | 7.58 |  | 1.36 (1.00, 1.73) | <0.001 |
| Wyoming | 4,735 | 0.82 |  | * | 3.85 |  | 3.03 (0.00, 6.04) | <0.001 |
| ^a^Per *All of Us* Data and Statistics Dissemination Policy, a participant count of 1 to 20 nor data/statistics that allow a participant count of 1 to 20 to be derived from other reported information cannot be published directly—thus indicated as *. | | | | | | | | |
| ^b^Positive value indicates overrepresentation in *All of Us*; negative value indicates underrepresentation in *All of Us*. | | | | | | | | |
| ^c^Calculated using the 2-sample test for equality of proportions | | | | |  |  |  |  |

| Table S5. Differences in non-Hispanic White representation between the US and *All of Us* | | | | | | | |  |
| --- | --- | --- | --- | --- | --- | --- | --- | --- |
|  | US | |  | *All of Us* | |  | Difference (95% CI), %^b^ |  |
| State | Count | Proportion, % | | Count | Proportion, % | | (*All of Us* - US) | *P* value^c^ |
| Alabama | 3,171,351 | 63.12 |  | 8,803 | 40.68 |  | -22.44 (-23.10, -21.79) | <0.001 |
| Alaska | 421,758 | 57.51 |  | 112 | 70.00 |  | 12.49 (5.39, 19.69) | <0.001 |
| Arizona | 3,816,547 | 53.37 |  | 25,209 | 56.72 |  | 3.35 (2.89, 3.81) | <0.001 |
| Arkansas | 2,063,550 | 68.52 |  | 273 | 65.31 |  | -3.21 (-7.77, 1.35) | 0.158 |
| California | 13,714,587 | 34.69 |  | 25,816 | 53.96 |  | 19.27 (18.83, 19.72) | <0.001 |
| Colorado | 3,760,663 | 65.13 |  | 1,266 | 79.08 |  | 13.94 (10.22, 14.20) | <0.001 |
| Connecticut | 2,279,232 | 63.21 |  | 1,289 | 52.31 |  | -10.89 (-12.87, -8.92) | <0.001 |
| Delaware | 579,851 | 58.57 |  | 110 | 78.57 |  | 20.00 (13.20, 26.80) | <0.001 |
| District of Columbia | 261,771 | 37.96 |  | 335 | 76.66 |  | 38.70 (34.73, 42.66) | <0.001 |
| Florida | 11,100,503 | 51.54 |  | 6,409 | 37.43 |  | -14.11 (-14.83, -13.38) | <0.001 |
| Georgia | 5,362,156 | 50.06 |  | 3,829 | 42.46 |  | -7.60 (-8.62, -6.58) | <0.001 |
| Hawaii | 314,365 | 21.60 |  | 116 | 63.04 |  | 41.44 (34.47, 48.42) | <0.001 |
| Idaho | 1,450,523 | 78.87 |  | 226 | 92.24 |  | 13.37 (10.02, 16.72) | <0.001 |
| Illinois | 7,472,751 | 58.32 |  | 10,920 | 33.46 |  | -24.86 (-25.38, -24.35) | <0.001 |
| Indiana | 5,121,004 | 75.47 |  | 920 | 79.24 |  | 3.77 (1.44, 6.11) | 0.003 |
| Iowa | 2,638,201 | 82.69 |  | 496 | 93.23 |  | 10.54 (8.41, 12.68) | <0.001 |
| Kansas | 2,122,575 | 72.25 |  | 862 | 87.51 |  | 15.26 (13.20, 17.33) | <0.001 |
| Kentucky | 3,664,764 | 81.33 |  | 354 | 90.54 |  | 9.20 (6.30, 12.10) | <0.001 |
| Louisiana | 2,596,702 | 55.75 |  | 1,226 | 31.56 |  | -24.19 (-25.65, -22.73) | <0.001 |
| Maine | 1,228,264 | 90.16 |  | 554 | 95.35 |  | 5.20 (3.48, 6.91) | <0.001 |
| Maryland | 2,913,782 | 47.17 |  | 1,382 | 75.15 |  | 27.98 (26.00, 29.96) | <0.001 |
| Massachusetts | 4,748,897 | 67.55 |  | 20,979 | 68.91 |  | 1.36 (0.84, 1.88) | <0.001 |
| Michigan | 7,295,651 | 72.40 |  | 12,877 | 72.28 |  | -0.11 (-0.77, 0.54) | 0.732 |
| Minnesota | 4,353,880 | 76.30 |  | 3,408 | 94.01 |  | 17.72 (16.94, 18.49) | <0.001 |
| Mississippi | 1,639,077 | 55.35 |  | 676 | 18.85 |  | -36.50 (-37.79, -35.22) | <0.001 |
| Missouri | 4,663,907 | 75.78 |  | 1,146 | 88.70 |  | 12.92 (11.20, 14.65) | <0.001 |
| Montana | 901,318 | 83.13 |  | 249 | 96.89 |  | 13.76 (11.63, 15.88) | <0.001 |
| Nebraska | 1,484,687 | 75.69 |  | 205 | 63.86 |  | -11.83 (-17.08, -6.57) | <0.001 |
| Nevada | 1,425,952 | 45.93 |  | 423 | 74.21 |  | 28.28 (24.69, 31.87) | <0.001 |
| New Hampshire | 1,200,649 | 87.16 |  | 873 | 94.28 |  | 7.12 (5.62, 8.61) | <0.001 |
| New Jersey | 4,816,381 | 51.85 |  | 1,299 | 63.68 |  | 11.83 (9.74, 13.91) | <0.001 |
| New Mexico | 772,952 | 36.50 |  | 466 | 69.04 |  | 32.53 (29.05, 36.02) | <0.001 |
| New York | 10,598,907 | 52.47 |  | 10,013 | 33.36 |  | -19.11 (-19.64, -18.58) | <0.001 |
| North Carolina | 6,312,148 | 60.46 |  | 1,155 | 76.80 |  | 16.33 (14.20, 18.46) | <0.001 |
| North Dakota | 636,160 | 81.65 |  | 285 | 92.83 |  | 11.18 (8.29, 14.07) | <0.001 |
| Ohio | 8,954,135 | 75.89 |  | 1,100 | 86.89 |  | 11.00 (9.14, 12.86) | <0.001 |
| Oklahoma | 2,407,188 | 60.80 |  | 256 | 83.66 |  | 22.86 (18.72, 27.01) | <0.001 |
| Oregon | 3,036,158 | 71.65 |  | 792 | 89.69 |  | 18.04 (16.03, 20.05) | <0.001 |
| Pennsylvania | 9,553,417 | 73.47 |  | 26,767 | 81.83 |  | 8.36 (7.94, 8.78) | <0.001 |
| Puerto Rico | 24,548 | 0.75 |  | 25 | 28.41 |  | 27.66 (18.24, 37.08) | <0.001 |
| Rhode Island | 754,050 | 68.71 |  | 439 | 89.96 |  | 21.25 (18.58, 23.91) | <0.001 |
| South Carolina | 3,178,552 | 62.10 |  | 692 | 30.52 |  | -31.58 (-33.47, -29.68) | <0.001 |
| South Dakota | 705,583 | 79.58 |  | 189 | 93.56 |  | 13.99 (10.60, 17.37) | <0.001 |
| Tennessee | 4,900,246 | 70.91 |  | 1,955 | 76.16 |  | 5.25 (3.60, 6.90) | <0.001 |
| Texas | 11,584,597 | 39.75 |  | 7,321 | 61.58 |  | 21.83 (20.96, 22.70) | <0.001 |
| Utah | 2,465,355 | 75.36 |  | 436 | 86.34 |  | 10.98 (7.98, 13.98) | <0.001 |
| Vermont | 573,201 | 89.13 |  | 183 | 97.86 |  | 8.73 (6.65, 10.80) | <0.001 |
| Virginia | 5,058,363 | 58.60 |  | 1,229 | 79.91 |  | 21.30 (19.30, 23.31) | <0.001 |
| Washington | 4,918,820 | 63.84 |  | 1,843 | 89.60 |  | 25.76 (24.44, 27.08) | <0.001 |
| West Virginia | 1,598,834 | 89.14 |  | 160 | 92.49 |  | 3.35 (-0.58, 7.28) | <0.001 |
| Wisconsin | 4,634,018 | 78.63 |  | 17,363 | 85.46 |  | 6.83 (6.34, 7.32) | <0.001 |
| Wyoming | 469,664 | 81.42 |  | 146 | 93.59 |  | 12.17 (8.33, 16.02) | <0.001 |
| ^a^Per *All of Us* Data and Statistics Dissemination Policy, a participant count of 1 to 20 nor data/statistics that allow a participant count of 1 to 20 to be derived from other reported information cannot be published directly—thus indicated as *. | | | | | | | | |
| ^b^Positive value indicates overrepresentation in *All of Us*; negative value indicates underrepresentation in *All of Us*. | | | | | | | | |
| ^c^Calculated using the 2-sample test for equality of proportions | | | | |  |  |  |  |

| Table S6. Differences in Other race/ethnicity representation between the US and *All of Us* | | | | | | | |  |
| --- | --- | --- | --- | --- | --- | --- | --- | --- |
|  | US | |  | *All of Us* | |  | Difference (95% CI), %^b^ |  |
| State | Count | Proportion, % | | Count | Proportion, % | | (*All of Us* - US) | *P* value^c^ |
| Alabama | 224,804 | 4.47 |  | 293 | 1.35 |  | -3.12 (-3.28, -2.97) | <0.001 |
| Alaska | 197,629 | 26.95 |  | * | 4.38 |  | -22.57 (-25.74, -19.40) | <0.001 |
| Arizona | 576,704 | 8.06 |  | 1,195 | 2.69 |  | -5.38 (-5.53, -5.22) | <0.001 |
| Arkansas | 190,033 | 6.31 |  | * | 1.91 |  | -4.40 (-5.71, -3.08) | <0.001 |
| California | 2,145,903 | 5.43 |  | 1,784 | 3.73 |  | -1.70 (-1.87, -1.53) | <0.001 |
| Colorado | 333,131 | 5.77 |  | 42 | 2.62 |  | -3.15 (-3.89, -2.33) | <0.001 |
| Connecticut | 172,023 | 4.77 |  | 57 | 2.31 |  | -2.46 (-3.05, -1.86) | <0.001 |
| Delaware | 50,449 | 5.10 |  | * | 2.14 |  | -2.95 (-5.35, -0.55) | 0.112 |
| District of Columbia | 34,864 | 5.06 |  | * | 3.43 |  | -1.62 (-3.33, 0.08) | 0.121 |
| Florida | 983,766 | 4.57 |  | 381 | 2.23 |  | -2.34 (-2.56, -2.12) | <0.001 |
| Georgia | 472,496 | 4.41 |  | 178 | 1.97 |  | -2.44 (-2.72, -2.15) | <0.001 |
| Hawaii | 448,548 | 30.82 |  | 27 | 14.67 |  | -16.15 (-21.26, -11.04) | <0.001 |
| Idaho | 108,355 | 5.89 |  | * | 1.22 |  | -4.67 (-6.04, -3.29) | 0.002 |
| Illinois | 479,455 | 3.74 |  | 662 | 2.03 |  | -1.71 (-1.87, -1.56) | <0.001 |
| Indiana | 306,182 | 4.51 |  | 25 | 2.15 |  | -2.36 (-3.19, -1.52) | <0.001 |
| Iowa | 131,844 | 4.13 |  | * | 0.94 |  | -3.19 (-4.01, -2.37) | <0.001 |
| Kansas | 184,125 | 6.27 |  | * | 0.61 |  | -5.66 (-6.14, -5.17) | <0.001 |
| Kentucky | 201,611 | 4.47 |  | * | 1.02 |  | -3.45 (-4.45, -2.45) | <0.001 |
| Louisiana | 200,750 | 4.31 |  | 66 | 1.70 |  | -2.61 (-3.02, -2.20) | <0.001 |
| Maine | 65,703 | 4.82 |  | * | 1.38 |  | -3.45 (-4.39, -2.50) | <0.001 |
| Maryland | 320,708 | 5.19 |  | 65 | 3.53 |  | -1.66 (-2.50, -0.81) | 0.001 |
| Massachusetts | 431,380 | 6.14 |  | 893 | 2.93 |  | -3.20 (-3.39, -3.01) | <0.001 |
| Michigan | 526,512 | 5.22 |  | 506 | 2.84 |  | -2.38 (-2.63, -2.14) | <0.001 |
| Minnesota | 316,664 | 5.55 |  | 45 | 1.24 |  | -4.31 (-4.67, -3.94) | <0.001 |
| Mississippi | 105,676 | 3.57 |  | 29 | 0.81 |  | -2.76 (-3.05, -2.47) | <0.001 |
| Missouri | 363,006 | 5.90 |  | 25 | 1.93 |  | -3.96 (-4.71, -3.21) | <0.001 |
| Montana | 124,554 | 11.49 |  | * | 0.39 |  | -11.10 (-11.86, -10.34) | <0.001 |
| Nebraska | 95,338 | 4.86 |  | * | 1.87 |  | -2.99 (-4.47, -1.51) | 0.013 |
| Nevada | 230,454 | 7.42 |  | 27 | 4.74 |  | -2.69 (-4.43, -0.94) | 0.014 |
| New Hampshire | 63,167 | 4.59 |  | * | 1.62 |  | -2.97 (-3.78, -2.15) | <0.001 |
| New Jersey | 372,975 | 4.02 |  | 72 | 3.53 |  | -0.49 (-1.13, 0.31) | 0.264 |
| New Mexico | 260,168 | 12.29 |  | * | 2.67 |  | -9.62 (-10.84, -8.40) | <0.001 |
| New York | 978,959 | 4.85 |  | 853 | 2.84 |  | -2.00 (-2.19, -1.82) | <0.001 |
| North Carolina | 561,059 | 5.37 |  | 35 | 2.33 |  | -3.05 (-3.81, -2.29) | <0.001 |
| North Dakota | 70,320 | 9.03 |  | * | 1.30 |  | -7.72 (-8.99, -6.45) | <0.001 |
| Ohio | 570,221 | 4.83 |  | 26 | 2.05 |  | -2.78 (-3.56, -2.00) | <0.001 |
| Oklahoma | 707,339 | 17.87 |  | * | 2.94 |  | -14.92 (-16.82, -13.03) | <0.001 |
| Oregon | 341,886 | 8.07 |  | 29 | 3.28 |  | -4.78 (-5.96, -3.61) | <0.001 |
| Pennsylvania | 524,016 | 4.03 |  | 641 | 1.96 |  | -2.07 (-2.22, -1.92) | <0.001 |
| Puerto Rico | 5,251 | 0.16 |  | * | 1.14 |  | 0.98 (-1.24, 3.19) | 0.022 |
| Rhode Island | 67,475 | 6.15 |  | * | 2.25 |  | -3.89 (-5.21, -2.58) | <0.001 |
| South Carolina | 228,610 | 4.47 |  | 20 | 0.88 |  | -3.58 (-3.97, -3.20) | <0.001 |
| South Dakota | 111,570 | 12.58 |  | * | 2.48 |  | -10.11 (-12.24, -7.96) | <0.001 |
| Tennessee | 313,333 | 4.53 |  | 44 | 1.71 |  | -2.82 (-3.32, -2.32) | <0.001 |
| Texas | 1,112,961 | 3.82 |  | 198 | 1.67 |  | -2.15 (-2.38, -1.92) | <0.001 |
| Utah | 197,539 | 6.04 |  | * | 2.97 |  | -3.07 (-4.55, -1.59) | 0.004 |
| Vermont | 34,266 | 5.33 |  | * | 1.60 |  | -3.72 (-5.53, -1.92) | 0.023 |
| Virginia | 475,579 | 5.51 |  | 45 | 2.93 |  | -2.58 (-3.43, -1.74) | <0.001 |
| Washington | 708,016 | 9.19 |  | 44 | 2.14 |  | -7.05 (-7.68, -6.42) | <0.001 |
| West Virginia | 80,403 | 4.48 |  | * | 1.16 |  | -3.33 (-4.92, -1.73) | 0.035 |
| Wisconsin | 271,635 | 4.61 |  | 246 | 1.21 |  | -3.40 (-3.55, -3.25) | <0.001 |
| Wyoming | 38,369 | 6.65 |  | 0 | 0.00 |  | -6.65 (-6.72, -6.59) | 0.001 |
| ^a^Per *All of Us* Data and Statistics Dissemination Policy, a participant count of 1 to 20 nor data/statistics that allow a participant count of 1 to 20 to be derived from other reported information cannot be published directly—thus indicated as *. | | | | | | | | |
| ^b^Positive value indicates overrepresentation in *All of Us*; negative value indicates underrepresentation in *All of Us*. | | | | | | | | |
| ^c^Calculated using the 2-sample test for equality of proportions | | | | |  |  |  |  |

| Table S7. Diversity of genetic ancestry within each self-reported racial and ethnic category in *All of Us* | | | | | | |  |  |  |  |
| --- | --- | --- | --- | --- | --- | --- | --- | --- | --- | --- |
|  | Self-reported race and ethnicity | | | |  |  |  |  |  |  |
| Race and ethnicity category | Hispanic or Latino | | Non-Hispanic Asian | | Non-Hispanic Black or African American | | Non-Hispanic White | | Other including multiracial | |
| Genetically-inferred ancestry |  |  |  |  |  |  |  |  |  |  |
| African | 1039 | (6.01%) | 36 | (1.16%) | 20,767 | (95.86%) | 304 | (0.58%) | 355 | (16.41%) |
| East Asian | * | (*) | 2049 | (65.95%) | * | (*) | * | (*) | 30 | (1.39%) |
| European | 306 | (1.77%) | * | (*) | * | (*) | 46,155 | (88.70%) | 544 | (25.15%) |
| Latino/Admixed American | 13,460 | (77.81%) | 51 | (1.64%) | 97 | (0.48%) | 1058 | (2.03%) | 179 | (8.28%) |
| Middle Eastern | * | (*) | * | (*) | * | (*) | * | (*) | 144 | (6.66%) |
| Other | 2488 | (14.38%) | 136 | (4.38%) | 781 | (3.61%) | 4480 | (8.61%) | 805 | (37.22%) |
| South Asian | * | (*) | 829 | (26.69%) | * | (*) | * | (*) | 106 | (4.90%) |
| ^a^All values are presented as count (%) within each self-reported race and ethnicity category (column total).  ^b^Genetic ancestry categories are consistent with the gnomAD, Human Genome Diversity Project, and 1000 Genomes classifications.  ^c^Per *All of Us* Data Statistics Dissemination Policy, a participant count of 1 to 20 nor data/statistics that allow a participant count of 1 to 20 to be derived from other reported information cannot be published directly—thus indicated as *. | | | | | | | | | | |

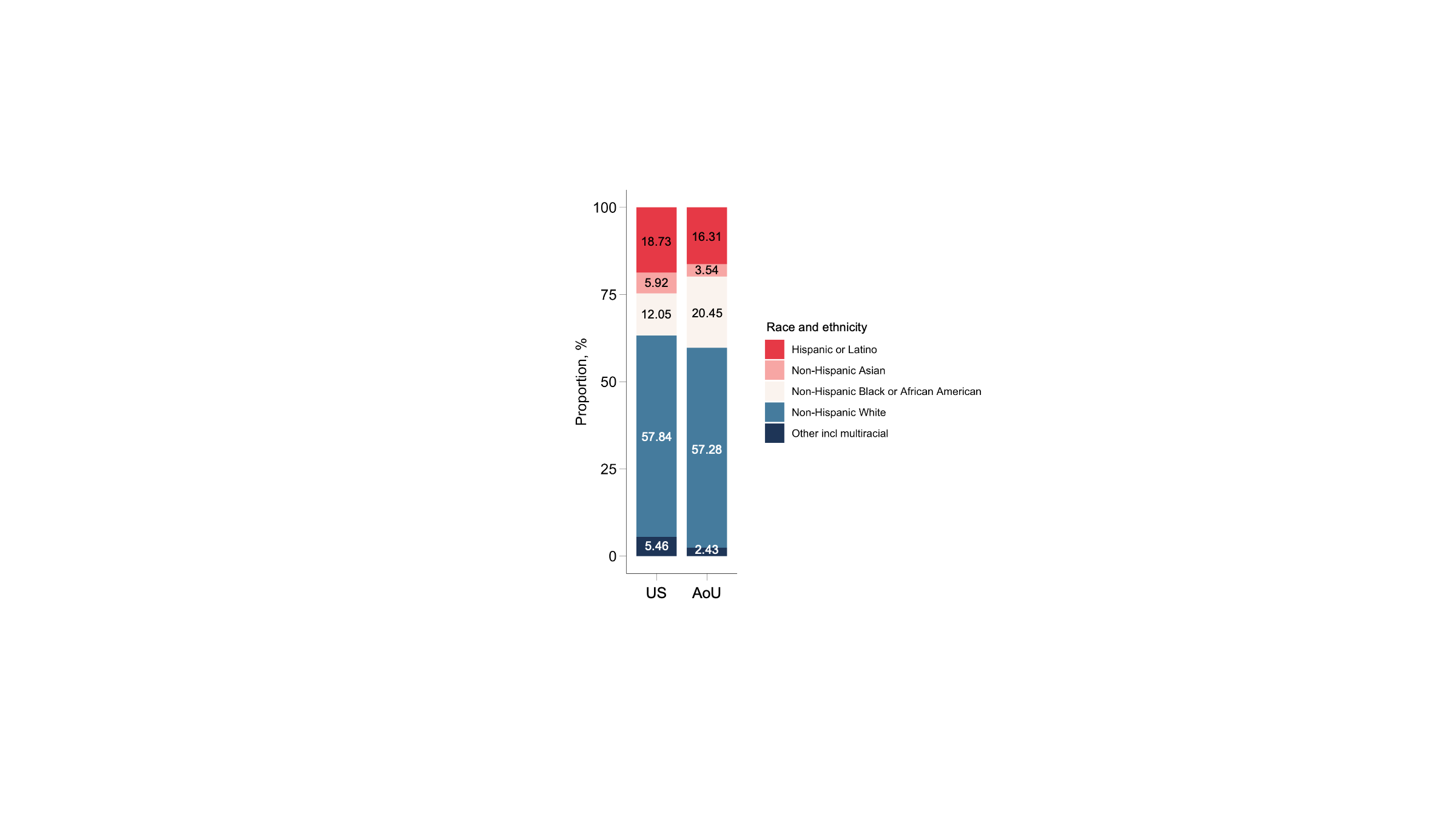

**Figure S1.** Comparison of racial and ethnic proportions between the US and AoU

^a^The US data on racial and ethnic distributions are projected by the American Community Survey 2020.

^b^Race and ethnicity are self-reported from fixed categories of Hispanic or Latino, Non-Hispanic Asian, Non-Hispanic Black or African American, Non-Hispanic White, and other including multiple race in accordance with the US Census Bureau scheme.

Abbreviation: AoU, *All of Us*

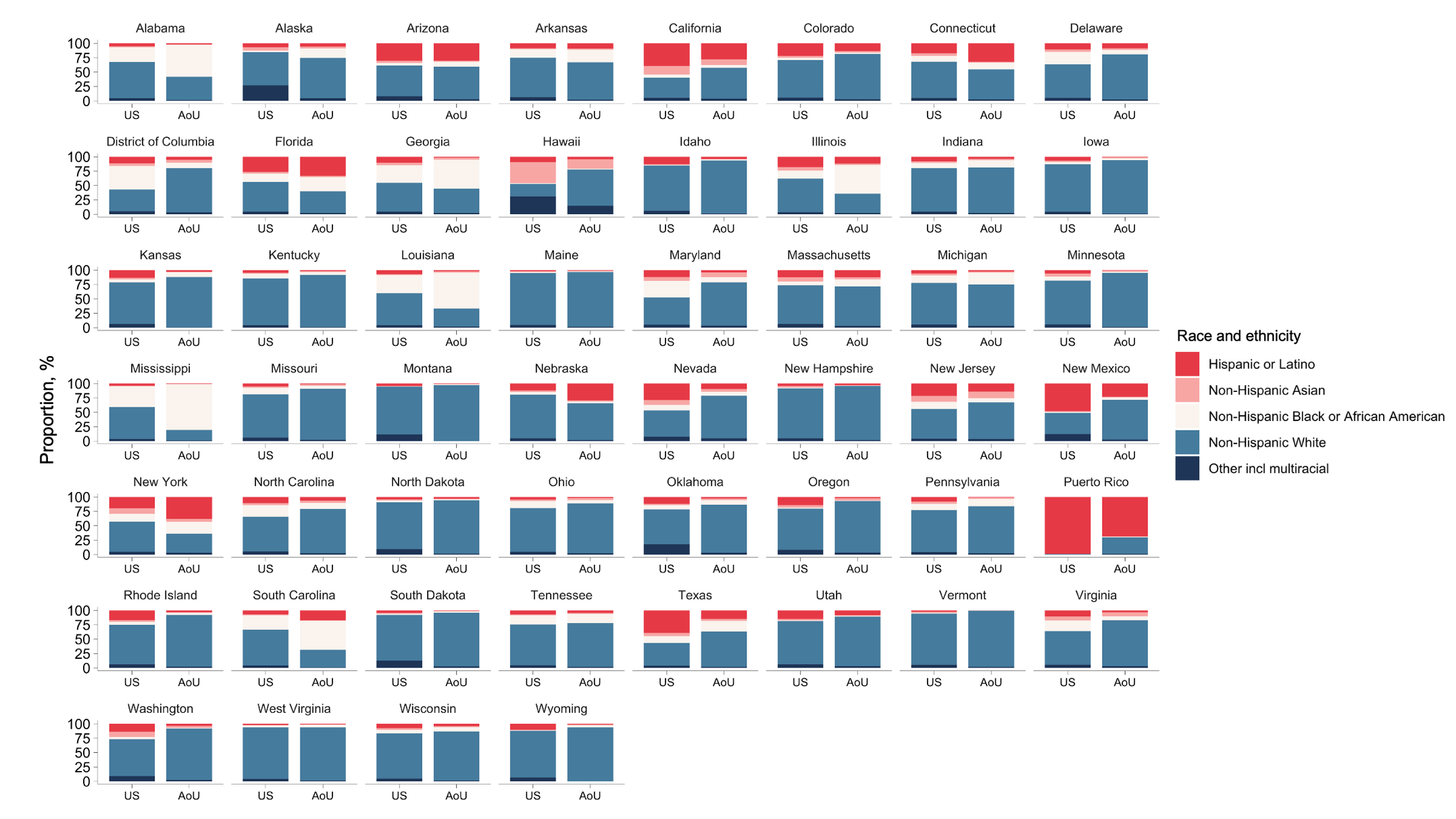

**Figure S2.** Comparison of racial and ethnic proportions between the US and AoU by state

^a^The US data on racial and ethnic distributions are projected by the American Community Survey 2020.

^b^Race and ethnicity are self-reported from fixed categories of Hispanic or Latino, Non-Hispanic Asian, Non-Hispanic Black or African American, Non-Hispanic White, and other including multiple race in accordance with the US Census Bureau scheme.

Abbreviation: Aou, *All of Us*

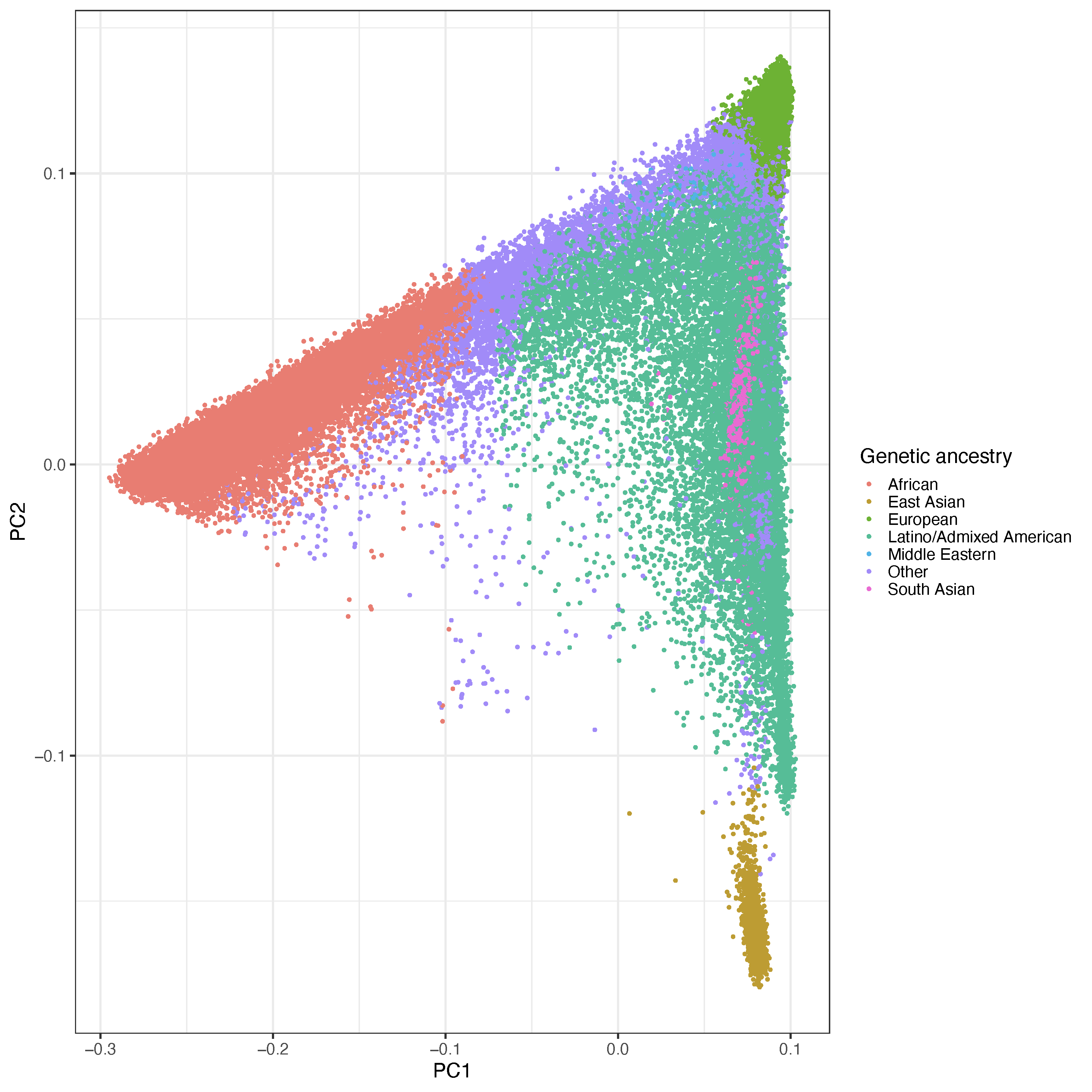

**Figure S3.** Principal component plot of *All of Us* participants

^a^Principal components of ancestry are displayed for 96,268 of 358,705 participants with whole genome sequencing data.

^b^Individuals (circle) are plotted by their first two genetic principal component coordinates and color-coded to reflect distinct genetically-inferred ancestry.

^c^Genetic ancestry categories are consistent with the gnomAD, Human Genome Diversity Project, and 1000 Genomes classifications.

^d^The “Other” category consists of individuals with predicted ancestry ≤90% by the random forest model.

Abbreviation: PC, principal component

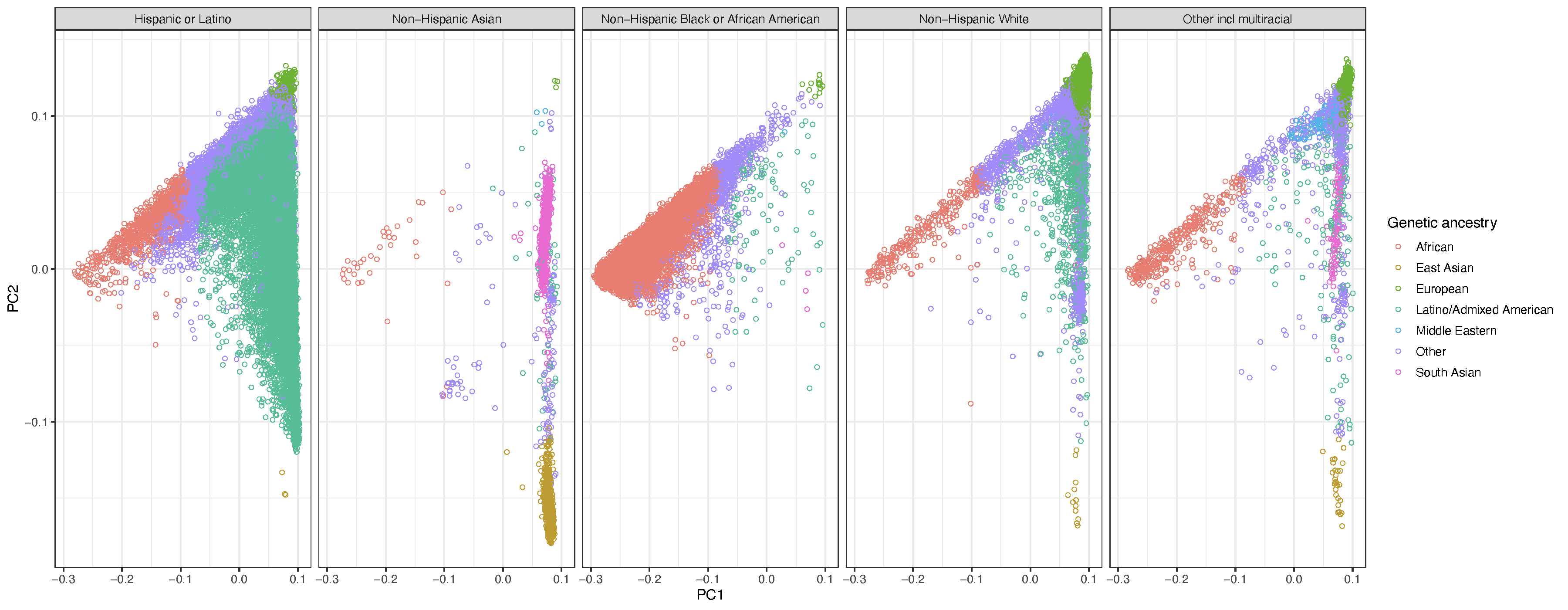

**Figure S4.** Heterogeneity in genetic ancestry within self-reported racial and ethnic category in *All of Us*

^a^Principal components of ancestry are displayed separately for individuals stratified by self-reported race and ethnicity. Color codings reflect genetically-inferred ancestry from 96,268 of 358,705 participants with whole genome sequencing data.

^b^Individuals (circle) are plotted by their first two genetic principal component coordinates.

^c^Genetic ancestry categories are consistent with the gnomAD, Human Genome Diversity Project, and 1000 Genomes classifications.

^d^The “Other” category consists of individuals with predicted ancestry ≤90% by the random forest model.

Abbreviation: PC, principal component
