## Supplementary material for "Representation of Race and Ethnicity in a Contemporary US Health Cohort: The *All of Us* Research Program": STAR Methods table

**KEY RESOURCES TABLE**

**Key resources table**

| Other | | |
| --- | --- | --- |
| Race and ethnicity | Concept ID^a^ | Subpopulations |
| Hispanic or Latino | 1586147 | Colombian, Cuban, Dominican, Ecuadorian, Honduran, Mexican, Puerto Rican, Salvadoran, Spanish, Unspecified |
| Non-Hispanic Asian | 1586142 | Asian Specific Indian, Cambodian, Chinese, Filipino, Hmong, Japanese, Korean, Pakistani, Vietnamese, Unspecified |
| Non-Hispanic Black or African American | 1586143 | African American, Barbadian, Caribbean, Ethiopian, Ghanian, Haitian, Jamaican, Liberian, Nigerian, Somali, South African, Unspecified |
| Non-Hispanic White | 1586146 | Dutch, English, European, French, German, Irish, Italian, Norwegian, Polish, Scottish, Spanish, Unspecified |
| Other | 903070 | Unspecified racial and ethnic subpopulations or individuals reporting multiple categories |

^a^Corresponds to value source concept identification number as indicated in the *All of Us* Researcher Workbench
